# Estimating the future global dose demand for Measles-Rubella microarray patches

**DOI:** 10.1101/2022.08.11.22278665

**Authors:** Melissa Ko, Stefano Malvoti, Thomas Cherian, Carsten Mantel, Robin Biellik, Courtney Jarrahian, Marion Menozzi-Arnaud, Jean-Pierre Amorij, Hans Christiansen, Mark Papania, Martin I. Meltzer, Balcha Girma Masresha, Desiree Pastor, David N. Durrheim, Birgitte Giersing, Mateusz Hasso-Agopsowicz

## Abstract

**Background:** Progress towards measles and rubella (MR) elimination has stagnated as countries are unable to reach the required 95% vaccine coverage. Microarray patches (MAPs) are anticipated to offer significant programmatic advantages to needle and syringe (N/S) presentation and increase MR vaccination coverage. A demand forecast analysis of the programmatic doses required (PDR) could accelerate MR-MAP development by informing the size and return of the investment required to manufacture MAPs.

**Methods:** Unconstrained global MR-MAP demand for 2030-2040 was estimated for three scenarios, for groups of countries with similar characteristics (archetypes), and four types of uses of MR-MAPs (use cases). The base scenario 1 assumed that MR-MAPs would replace a share of MR doses delivered by N/S, and that MAPs can reach a proportion of previously unimmunised populations. Scenario 2 assumed that MR-MAPs would be piloted in selected countries in each region of the World Health Organization (WHO); and scenario 3 explored introduction of MR-MAPs earlier in countries with the lowest measles vaccine coverage and highest MR disease burden.

**Results:** For the base scenario (1), the estimated global PDR for MR-MAPs was forecasted at 30 million doses in 2030 and increased to 220 million doses by 2040. Compared to scenario 1, scenario 2 resulted in an overall decrease in PDR of 18%, and scenario 3 resulted in a 21% increase in PDR between 2030-2040.

**Conclusions:** Significant demand is expected for MR-MAPs between 2030-2040, however, efforts are required to address remaining data quality, uncertainties and gaps that underpin the assumptions in this analysis.

**Key points:** the delivery of measles and rubella vaccines with microarray patches (MR-MAPs) could disrupt the immunization landscape. We estimated the demand for MR-MAPs between 2030-2040 at 4.05 billion doses. This analysis will inform the size of investment required to manufacture MR-MAPs.

## Introduction

Prior to widespread vaccination, major epidemics of measles occurred every two to three years and measles caused an estimated 2.6 million deaths globally each year, while four babies for every 1,000 live births worldwide were born with congenital rubella syndrome (CRS).[1, 2] Measles and rubella remain a major cause of worldwide morbidity and mortality with an estimated 7.5 million measles cases and more than 60,700 measles-related deaths in 2020, and around 100,000 infants born with CRS each year.[1, 3] To eliminate transmission and prevent outbreaks of measles and rubella at least 90-95% of all children must receive both measles and rubella first (MCV1) and second vaccine doses (MCV2)[4].

While the global coverage of MCV1 increased from 72% to 84% between 2000 and 2020, it has stagnated at around 83-85% for the past 10 years.[5] MCV2 coverage has been steadily increasing as countries introduce the second dose into their routine schedules but has now also plateaued at around 70%.[5] Furthermore, countries and regions are struggling not only to achieve but sustain their measles elimination status through high and widespread levels of vaccination. [6] The impact of the SARS-CoV-2 pandemic and its disruptions to routine immunization services and supplemental immunization activities (SIA) have highlighted the precarious situation with regards to measles control and elimination, with the reduction in global MCV1 coverage likely to ‘fuel a resurgence of measles’.[5, 7, 8]

Efforts to increase coverage and reduce measles and rubella burden could be assisted by the application of innovative technologies such as microarray patches (MAPs) to help reach those communities that are currently under-immunized or completely missed by vaccination (un-immunized). A microarray patch (MAP) consists of hundreds to thousands of tiny projections that deliver vaccine just below the skin surface. Measles-Rubella MAPs (MR-MAPs) are anticipated to be easier to administer than needle-and-syringe (N/S) and be less burdensome on vaccinators and the immunization system, given that they would not require reconstitution, they come as a single dose presentation, have the potential for increased thermostability, and reduced weight. Further the needleless presentation could address some vaccine hesitancy due to an increasing number of painful injections administered during an immunization session. MAP characteristics required for maximum impact in low- and middle-income countries (LMICs) have been described in the MR-MAP Target Product Profile.[9] Given these product characteristics, MR-MAPs are anticipated to be delivered by less trained personal and together with the attributes noted above, facilitate reaching the un-(zero-dose) and under-immunized populations by minimizing delivery challenges. This improved reach is anticipated to ultimately contribute to increasing MCV1 and MCV2 coverage and achieving MR elimination goals. Previous studies have demonstrated MAP acceptability to care givers, its suitability for paediatric use, and preference over N/S administration.[10, 11]

While two MR-MAP products have entered Phase I clinical development in 2021 (NCT04394689 and ACTRN12621000820808), they continue to face several hurdles impeding development and slowing licensure timelines. The path to commercialization and uptake is critical since MR-MAP developers or their commercialization partners will need to invest in building and validating MAP manufacturing facilities, and this has not yet been achieved for any vaccine-MAP. Programmatic Doses Required (PDR) are defined as the average estimated number of doses required to meet immunization program needs, whether these are routine or campaign. The PDR includes wastage, which is dependent on the vaccine presentation, and buffer stock. Clarity on the potential demand of PDR and how countries plan to use MR-MAPs could help to inform manufacturing investment decisions. A recent analysis estimates that MR-MAPs could be available to all countries by 2030.[12]

This article summarises the demand estimates for MR-MAPs between 2030-2040. It aims to inform analyses such as cost-effectiveness, willingness to pay, size of manufacturing investment required to manufacture MR-MAPs; as well as other decisions like preparation of appropriate policies, communication or training material to allow for rapid MR-MAP introduction.

## Methodology

To forecast the global MR-MAP demand from 2030-40, a five-step approach was used. This forecast was unconstrained from programmatic, supply, and price perspectives. The methodology described here is a high level, non-technical summary of the approach. A detailed summary of the methodology can be found in Annex A, together with an Excel file “annex.xls” that contains data, sources, all assumptions used in this analysis, and a model to estimate the year of introduction of MR-MAPs. Additional sensitivity analyses can be found in Annex B.

First, **a demand expressed in PDR was estimated for measles-containing vaccines (MCV) for the years 2030-2040** by leveraging the methodology of the WHO’s Market Information for Access to Vaccine’s (MI4A) MCV Global Market Study, which contains global MR PDR estimates up to 2030.[13] MI4A is a study that provides a global perspective on vaccine markets, including for measles monovalent, MR, and Measles, Mumps, and Rubella (MMR) or Measles, Mumps, Rubella, and Varicella (MMRV), until 2030 using a population-based methodology, with protocols and assumptions established and reviewed by experts.[13] Briefly, the size of the target population, MCV1 and MCV2 coverage, frequency of SIAs, and buffer and stock wastage in 2030-2040 were used to estimate the MCV PDR demand for these years (annex A, step 1).

Second, **estimates of the number of N/S doses calculated in step 1 that would be replaced by MR-MAPs** were made. To achieve this, a set of criteria were identified to predict the year in which a country would adopt MR-MAPs. Then, a market penetration rate of MR-MAPs was applied. Market penetration is defined as the percentage of PDR utilizing MR-MAPs and assumes that MR-MAPs are delivered alongside the N/S presentation. Countries with similar patterns of MR use, or location, were grouped into four archetypes and 16 key countries (text box 1; annex A, step 2).

### Text box 1: Country archetypes

**A:** Countries exclusively using MMR or MMRV

**B:** Countries using MMR or MMRV in routine schedules but utilizing Measles monovalent and MR for SIA activities

**C:** Countries using MR located in WHO’s African & Eastern Mediterranean Region

**D:** Countries that use M or MR in WHO’s Southeast Asian & Western Pacific Region

**16 Key Countries:** Afghanistan, Bangladesh, Brazil, Chad, Democratic Republic of the Congo, Ethiopia, India, Indonesia, Mozambique, Nigeria, Pakistan, Philippines, South Africa, Tanzania, Uganda, United States of America

Third, **additional PDR was estimated given the anticipated increased reach of MR-MAPs** to zero-dose or under-immunized populations, including remote rural, security compromised, urban slums and missed opportunity for vaccination (MOV) populations. This included defining and developing a set of assumptions concerning definitions, size and vaccine coverage of these additional populations, recommended buffer stocks, and anticipated wastage rates (annex A, step 3).

Fourth, to understand who would administer MR-MAPs and where, **the total MR-MAP PDR from steps 2 and 3 was allocated to the previously defined MR-MAP Use Cases (UCs)** using the dimensions of delivery location and service provider (text box 2). The UCs were developed through consultations with National Immunization Programme managers as well as regional and national WHO focal points, and through desk reviews of published and unpublished literature on their potential feasibility and acceptability. These UCs were leveraged to develop an initial estimate of MR-MAP PDR from 2030-40. This exercise estimated the global PDR only for MR-MAPs delivered in a fixed health post or through outreach with or without cold chain capacity (UC1, UC2, and UC3/4), while additional research is currently being conducted to estimate PDR for self-administered MR-MAPs (UC5 and UC6), (annex A, step 4).

### Text box 2: Description of the MR-MAP Use Cases

UC1: Delivery in a fixed health post with full cold chain capabilities by health worker or community health worker.

UC2: Outreach delivery with reduced cold chain capabilities or no cold chain by health worker.

UC3: Outreach delivery with reduced cold chain capacities by community health worker.

UC4: Outreach delivery with no cold chain by community health worker in their ‘home’ community.

UC5: Self-administration with health worker or community health worker supervision.

UC6: Self-administration without health worker or community health worker supervision, but may include supervision by another individual (e.g., school teacher, community leader, etc). for monitoring purposes.

Finally, given the level of uncertainty for key assumptions that could significantly impact MR-MAP PDR, three scenarios were explored. The base scenario 1 assumed that MR-MAPs would replace a share of MR doses delivered by N/S, and that MAPs could reach a proportion of previously unimmunised populations. Scenario 2 assumed that MR-MAPs would be first piloted in selected countries in each WHO region prior to wider scale use in countries; and scenario 3 explored earlier introduction of MR-MAPs in countries with the lowest measles vaccine coverage and highest MR disease burden (annex A, step 5).

Throughout the development of the demand forecast, the Working Group of Experts on MR-MAPs, the MI4A Advisory Group and key country experts were consulted on the methodology and assumptions utilized.

Figure 1 provides an overview of the five-step process.

**Figure 1:**
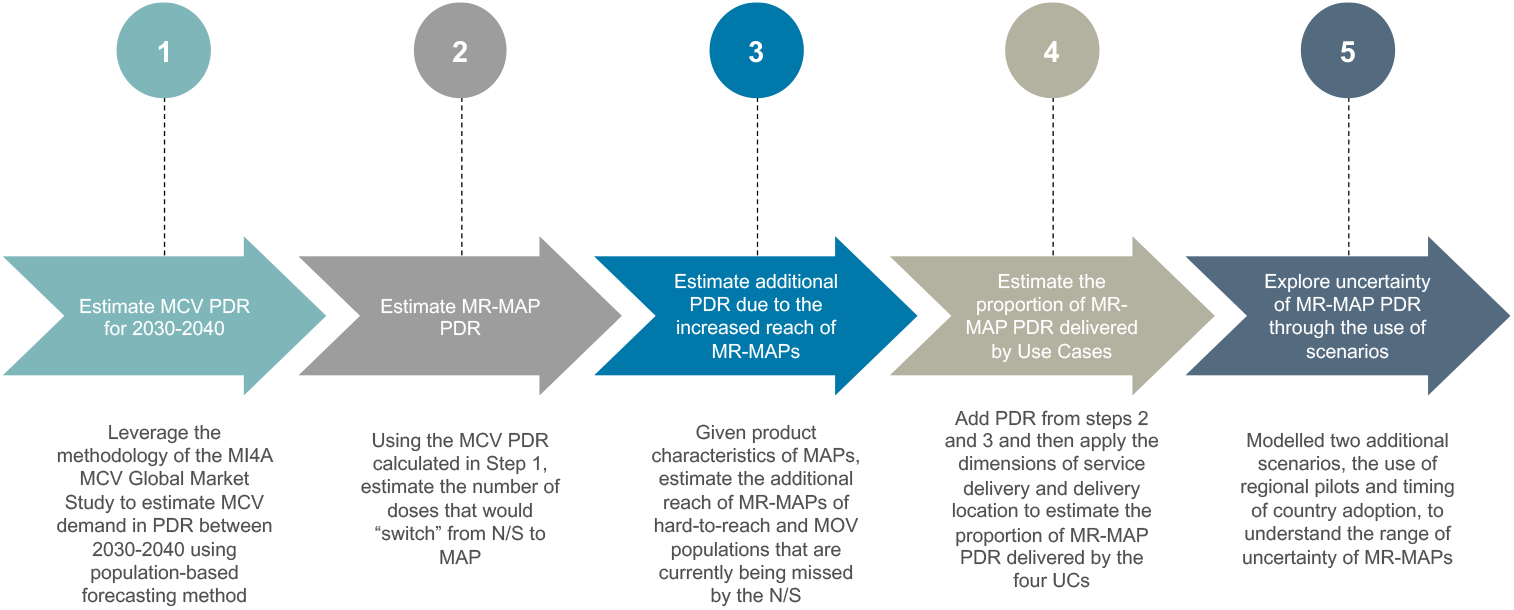
Overview of the four-step approach to develop MR-MAP demand forecast. MR-MAP: measles and rubella microarray patches; MCV: measles containing vaccines; PDR: programmatic doses required; N/S: needle and syringe; MOV: missed opportunities for vaccination; UCs: use cases.

## Results

### Scenario 1 (Base) results

All numbers presented in this section are approximate. We estimated the PDR for MCV between 2030-2040 at 4.05 billion doses, with 3.05 billion doses delivered through routine immunization, and ∼1 billion doses delivered through SIAs. After applying additional steps including country adoption and market penetration rates (see methods), we estimated the PDR for MR-MAPs between 2030-2040 at 1.46 billion doses. We calculated the additional MR-MAP PDR to reach populations not receiving MCV vaccines due to MOV or living in hard-to-reach areas (urban slums, remote rural, and security compromised populations) at 0.25 billion doses between 2030-2040. This resulted in a total PDR for MR-MAPs between 2030-40 of 1.71 billion doses.

The initial MR-MAP PDR in 2030 is estimated at 30 million doses, expected to peak in 2036 at 250 million doses, and level out at 220 million doses in 2040. We estimated that MAPs will deliver a total of 9% (30 million doses) MR PDR in 2030, and steadily increase to approximately 76% (220 million doses) by 2040.[13] Figure 2 provides an overview of the total number of MR PDR delivered by MAPs and N/S in countries forecasted to use MR in their national immunization schedules.

**Figure 2:**
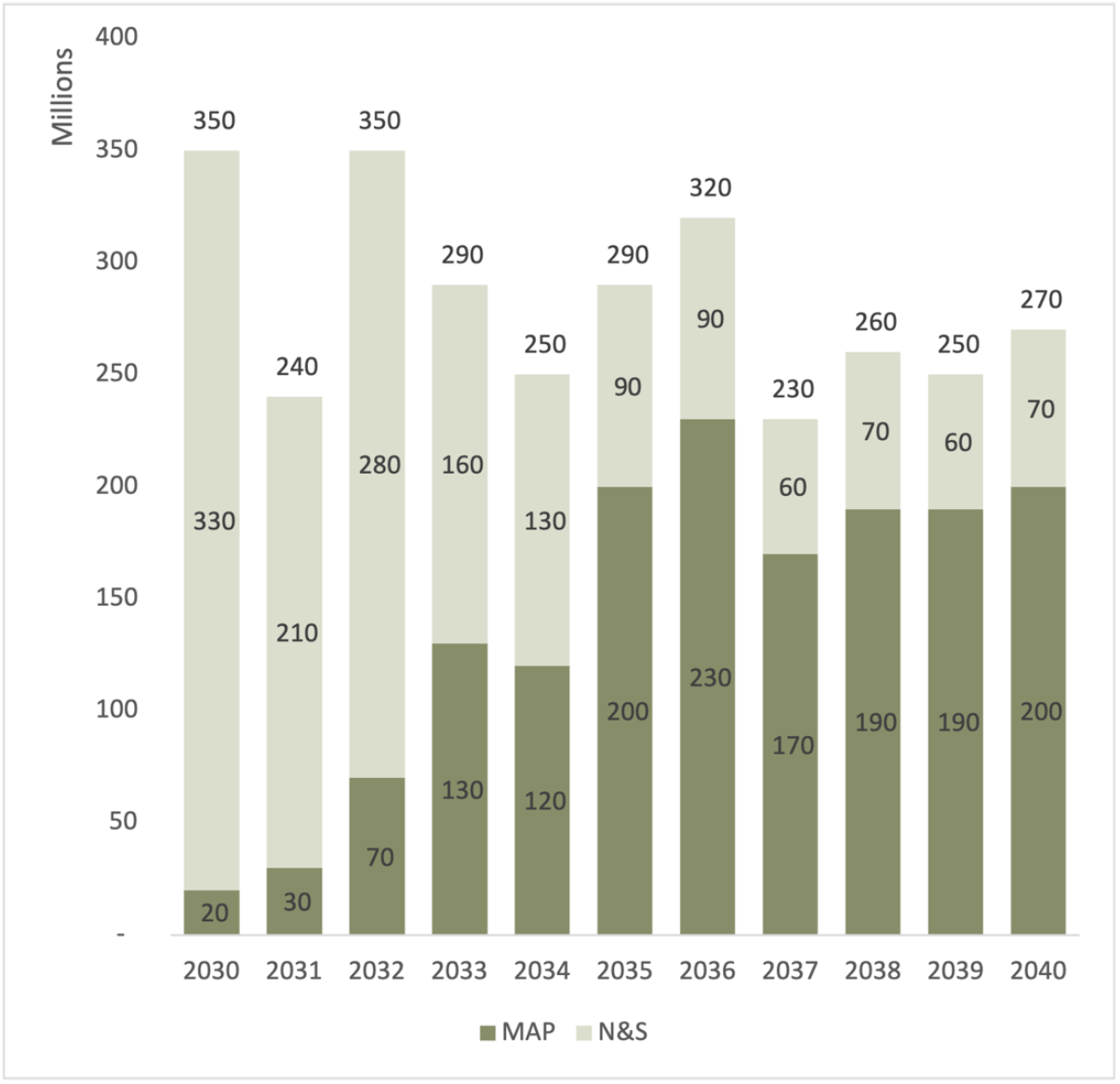
Estimated MR PDR delivered by MAPs and N/S for countries using MR in their national immunization schedules, between 2030-2040. MR: measles rubella; PDR: programmatic doses required; MAPs: microarray patches; N/S: needle and syringe.

The majority of the 2030 estimated MR-MAP PDR is from 16 key countries (37%) and Group A countries (50%) with Group B, C, and D accounting for 12% of total PDR. However, by 2040, the percentages are estimated to shift with the 16 key countries accounting for the largest share (61%) of total global PDR, followed by countries in Group C (27%) and countries in Group A, B, and D accounting for 3%, 5% and 4%, respectively.

Figure 3 and table S1 in Annex C provide an overview of the estimated global MR-MAP PDR by country group.

**Figure 3:**
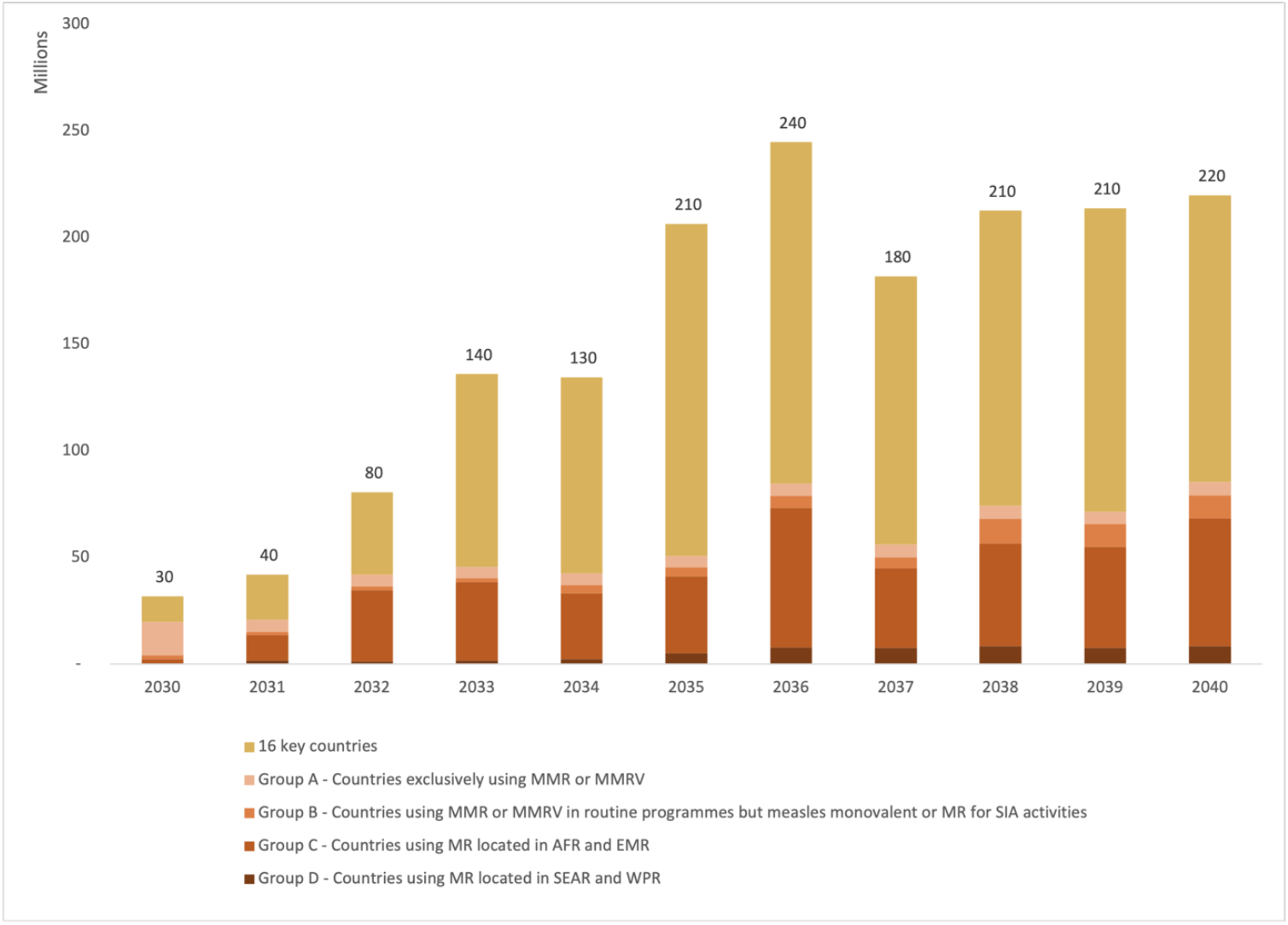
Estimated Global MR-MAP PDR – by country group, between 2030-2040. MR: measles and rubella; MAP: microarray patches; PDR: programmatic doses required; SEAR: WHO South East Asia Region; WPR: WHO Western Pacific Region; AFR: WHO African Region; EMR: WHO Eastern Mediterranean Region; MMR: measles, mumps, rubella; MMRV: measles, mumps, rubella, varicella; SIA: supplementary immunization activities.

In 2030, 61% of MR-MAP PDR is forecasted to be delivered at fixed health posts with full cold chain capabilities by health workers or community health workers (UC1). The remaining 39% of MR-MAP PDR is anticipated to be used as part of outreach activities or UC2 and UC3/4, i.e., when there is limited or no cold chain availability and MAPs will be administered by health workers or community health workers. By 2040, 52% of MR-MAP PDR is forecasted to be used in UC1 and 47% in UC2 and UC3/4, showing a slight shift in how doses may be used once most countries have adopted MR-MAPs. Figure 4 and table S2 in Annex C provide an overview of the estimated MR-MAP PDR by UC.

**Figure 4:**
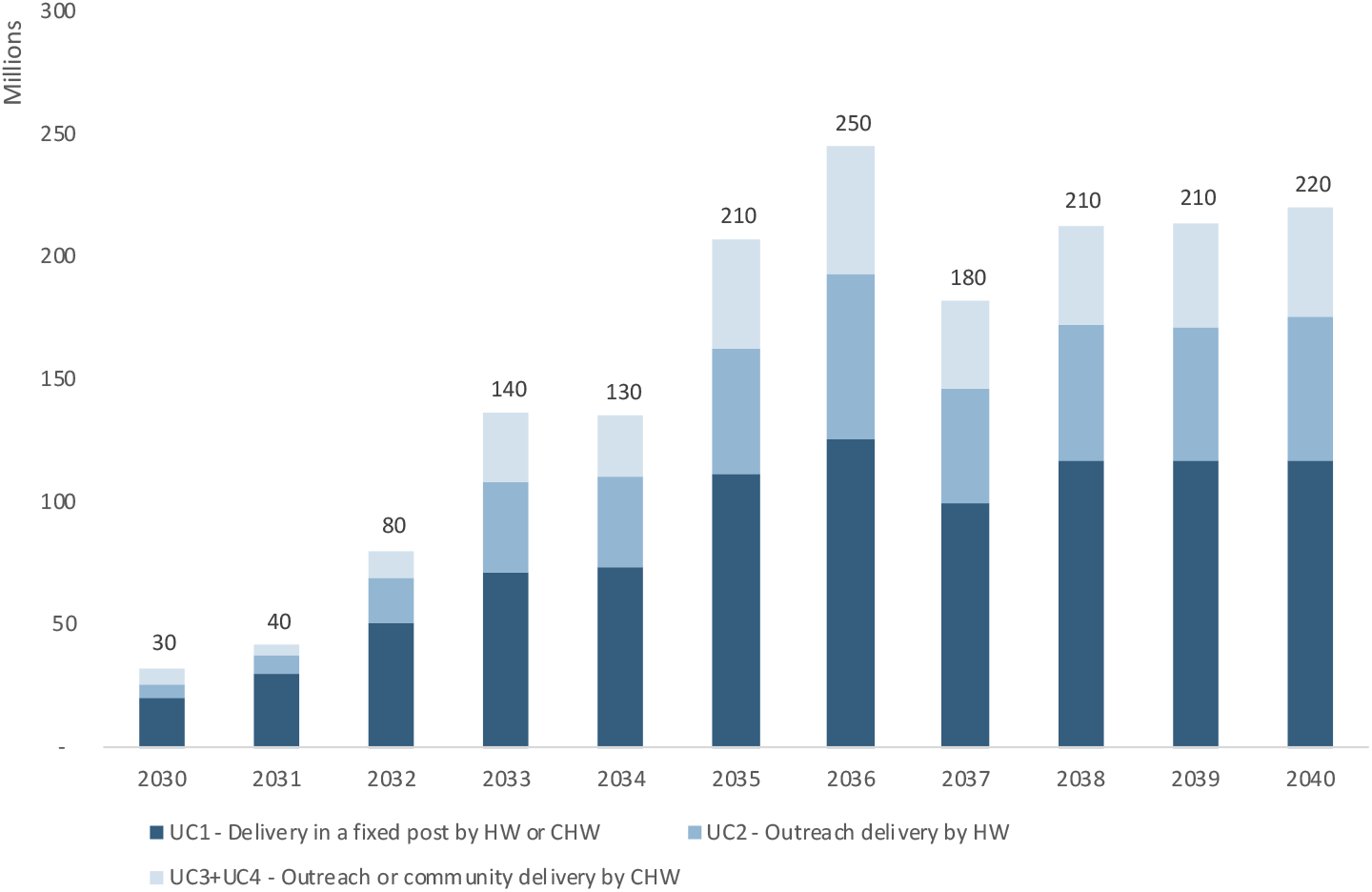
Estimated global PDR for MR-MAPs from 2030 to 2040 by UC. PDR: programmatic doses required; MR: measles and rubella; MAPs: microarray patches; UC: use case; HW: health worker; CHW: community health worker.

### Exploring uncertainty of global MR-MAP demand with scenarios

In addition to scenario 1 (baseline), two other scenarios were developed to investigate the uncertainty in the trajectory of MR-MAP demand between 2030 and 2040.

Scenario 2 explores the use of **regional MR-MAP pilots** and results in lower MR-MAP PDR estimates (130 million doses) over the first five years compared to the 290 million doses in Scenario 1 (Base) and in a 18% reduction of MR-MAP PDR over the 2030-40 period.

Scenario 3 explores more **accelerated MR-MAP adoption in countries with the greatest needs** and estimates an increase in PDR of 186% (830 million doses) in the first five years and an overall increase of 21% in PDR over the 2030-40 period compared to Scenario 1.

While the MR-MAPs PDR estimates for each of the scenarios vary, particularly in the first half of the forecasting period, the PDR estimates ultimately converge towards the latter part of the forecasting period. Figure 5 and table S3 in Annex C provide an overview of the estimated MR-MAP PDR by scenario.

**Figure 5:**
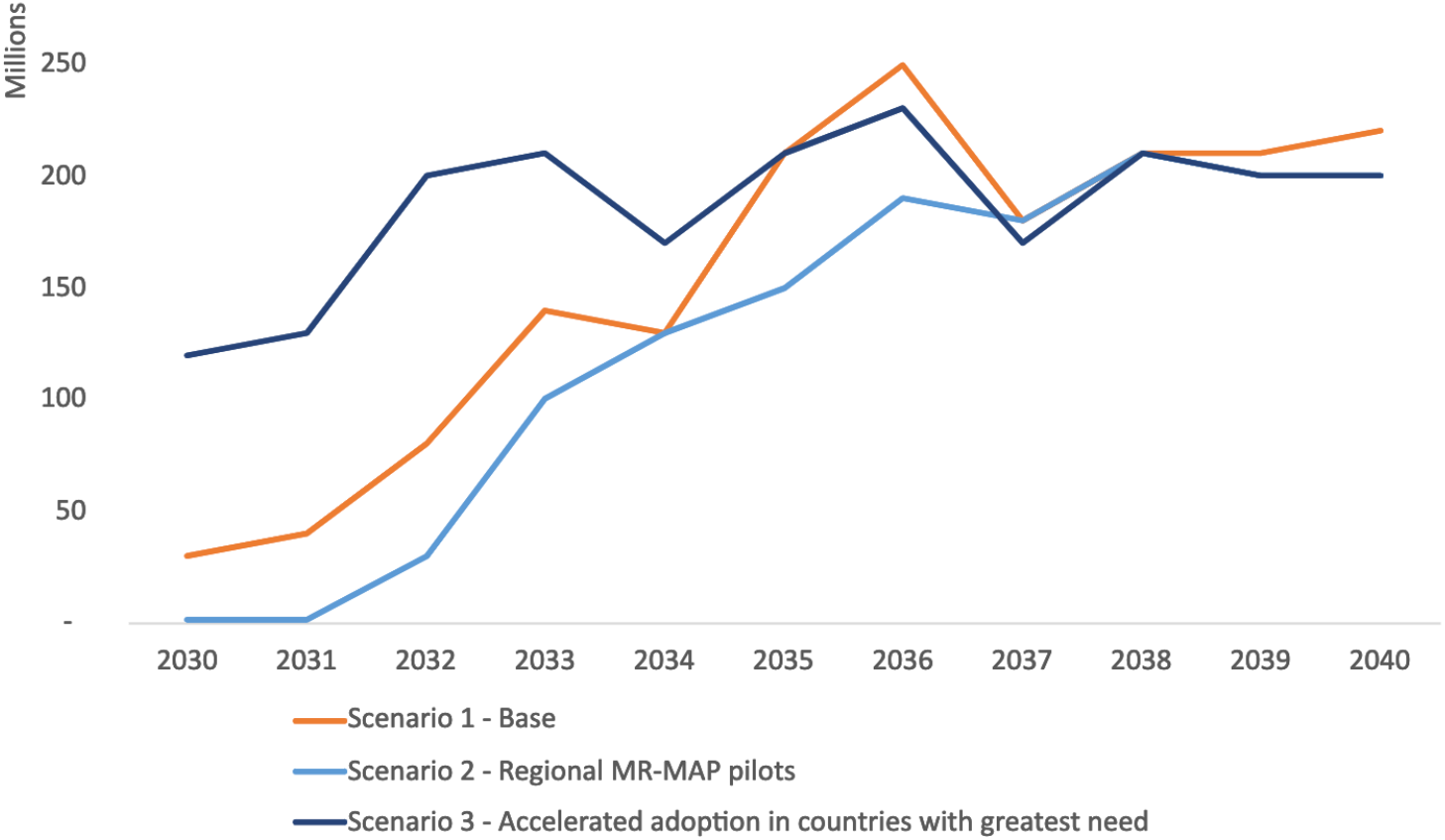
Comparison of global MR-MAP PDR by scenario, between 2030-2040. PDR: programmatic doses required; MR: measles and rubella; MAPs: microarray patches.

### Limitations

The limitations of this unconstrained demand forecast are largely driven by the limited evidence and data to inform key assumptions, including on how those data may evolve over the next 20 years. We conducted sensitivity analyses to measure the impact of data uncertainty on the estimates of MR-MAP demand between 2030-2040 and report the results in annex B.

In particular the data estimating the size of hard-to-reach populations as well as those prone to MOV that could be reached by using MR-MAPs was scarce with substantial uncertainty in how the situation will evolve over the next 20 years. However, sensitivity analyses revealed that even major changes in the assumptions of estimated population size (e.g., a decrease by 20%) only had a minor impact on the global PDR estimates with a decrease of 1 million doses (see supplemental Annex B for additional information on the methodology and results of the sensitivity analyses). In contrast, the sensitivity analyses revealed that assumptions around the anticipated reach or coverage of MR-MAPs, particularly in the hard-to-reach and MOV populations, and the market penetration of MR-MAPs significantly impacted the estimated PDR. These assumptions are currently being driven by limited data up to 2020, and gaps were filled by expert opinion and extrapolations made to 2040. Further revisions will require research and consultation to generate a stronger evidence-base.

Other assumptions relating to the delivery of MR-MAPs by less trained individuals (e.g., community health workers) may not be valid due to prevailing and future legal and policy constraints in some countries. These assumptions could also considerably impact global PDR, particularly the split of PDR between the different UCs. Additional evidence is required to understand if and how lesser trained individuals could administer MR-MAPs.

Finally, all assumptions are subject to the perceived benefit, operational feasibility, and cost-effectiveness of switching to MR-MAPs in different country situations. We have assumed that MR-MAPs will have desirable product characteristics based on the Target Product Profile. [9] However, uncertainties remain (e.g., thermostability, wear time, vaccine price, ability to reach zero-dose and under-immunized, and impact on cold chain) and these characteristics will likely contribute to country decision making on the adoption and use of MR-MAPs.

As MR-MAPs proceed through the clinical trial phases and the product attributes and operational feasibility become clearer, it will be important to re-visit the applied assumptions and update the demand forecast.

## Discussion

This work represents a first attempt to estimate the unconstrained global PDR of MR-MAPs while considering potential UCs, to inform global discussions and decision making about the investment in, development, introduction, and use of MR-MAPs. As additional information becomes available, the current assumptions should be adjusted to improve the accuracy of the estimates.

We estimate that MR-MAPs PDR will stabilize by 2038 at approximately 210 million doses per year and will account for approximately 76% of total MR PDR. In 2040, approximately 53%, 27%, and 20% of total MR-MAP PDR will be contributed by UC1, UC2, and UC3/4, respectively (Figure 4). This demand forecast indicates that there will be a substantial and sustainable role for MR-MAPs as part of national immunization programmes. Finally, we estimate that a significant portion of MR-MAP PDR will be used as part of routine immunization (73%) compared to SIAs (27%) by 2040.

Given that MR-MAPs are a new vaccine presentation, a pilot implementation strategy may be the most realistic scenario for their roll-out (Scenario 2). If designed appropriately this strategy could provide valuable evidence on vaccine acceptability and feasibility of uptake and identify best practices to assist countries with their decision making while demonstrating and communicating the benefits of MR-MAPs. Pilot strategies have previously proven useful for countries with the introduction of vaccines using different immunization strategies and varying delivery costs a (e.g., Human papillomavirus (HPV) vaccination demonstration projects and malaria vaccine pilots).

Scenario 3, which forecasts the use of MR-MAPs in countries with the greatest needs, reflects the global public health community’s ambition to urgently utilize MR-MAPs as a critical tool to overcome the limitations of current N/S presentations and close coverage gaps. This scenario highlights the potential role that MR-MAPs could play in achieving MR elimination goals.

Additional data and evidence will need to be gathered to refine these estimates, particularly exploring how countries are making decisions on the adoption of MR-MAPs, and how quickly countries could adopt MR-MAPs. Finally, this demand forecast was considered independent of price assumptions to investigate the full potential of demand, which can ultimately impact Cost of Goods Sold and price. There is a need to further explore how a potential increase in price will be off-set by cost savings in the immunization system and a potentially increased health and economic impact, including the ability of MR-MAPs to reach zero-dose and under-immunized children, which would result in reduced morbidity and mortality. This could play an important role in informing decisions by countries on adopting MR-MAPs.

## Conclusions

While MR-MAPs have recently entered Phase I clinical trials, questions on their anticipated use and impact remain (text box 3), which could influence the demand forecast and ultimately affect investment decisions, clinical development strategies and timelines. The use of MAP presentations could potentially change the immunization landscape by addressing some of the current barriers that countries face related to zero-dose and under-immunized children. Thus, it is imperative that these outstanding issues are addressed through open dialogue with countries and manufacturers. As these issues are resolved, the global demand forecasting methodology and assumptions for MR-MAPs will need to be continuously revised to reflect the latest data and information. This can improve the accuracy and reliability of the estimates of MR-MAP demand, which can play an important role in providing confidence to manufacturers of their investments.

### Text box 3: Key questions that can impact the demand forecast

- What is the anticipated reach or coverage of MR-MAPs in hard-to-reach and MOV populations?
- What is the market penetration of MR-MAPs and whether they will replace the N/S presentation?
- Can MR-MAPs can be delivered by lesser trained individuals?
- What is the quantification of the perceived benefit, operational feasibility, and cost effectiveness of using MR-MAP in different country situations and contexts?
- Will a potential price increase be offset by other cost savings related to cost savings in logistics, transportation, use of lesser trained individuals, etc?

## Supporting information

Supplemental Annex

Supplemental Annex- Data sets

## Data Availability

All data produced in the present work are contained in the manuscript

## Funding

This work was supported by Bill & Melinda Gates Foundation grant to World Health Organization [INV-005318].

## Acknowledgements

The authors thank the Working Group of Experts on MR-MAPs and MI4A Advisory Group who provided invaluable guidance and feedback on the development of the methodology and scenarios. The authors acknowledge immunization experts in countries who shared their expertise and feedback to review the methodology and develop country or regional assumptions.

## Supplemental Annex A: Additional information on the methodology and assumptions

### Step 1: Estimate MCV PDR for 2030-2040

#### Estimate the size of the target population, its characteristics, and immunization strategies

##### Data: annex.xls, sheets 2a-2g

To calculate the size of the future target populations, and the MCV PDR for 2030-2040, the medium estimates for the number of surviving infants and nine to 59 month old children were obtained from the United Nations World Population Prospectus (WPP) for years 2030-2040, which provides population estimates for 183 of the 194 UN countries.[13] Two delivery strategies were considered: (i) two MCV doses delivered in routine immunization at nine to 12 months and at 15-18 months of age; and (ii) one MCV dose delivered as part of supplemental immunization activities (SIAs) to children aged nine to 59 months of age every two to five years until optimal immunity in the target population can be achieved.[4, 14, 15] We assume that all countries in the analysis will have introduced MCV2 by 2030 (e.g., N/S introduction date).[13]

#### Forecast MCV1 and MCV2 coverage estimates

##### Data: annex.xls, sheets 4a-4b

The changes in MCV1 and MCV2 coverage between 2030 and 2040 were estimated using the 2019 WHO/UNICEF Estimates of National Immunization Coverage (WUENIC) MCV coverage estimates, leveraging the MI4A methodology, and based on the following assumptions: (i) if coverage is less than 70%, then an annual growth of 3% would be applied; (ii) if coverage was between 70-85% then an annual growth of 1% would be applied; and (iii) finally if coverage was greater than 85%, then an annual growth of 0.5% would be applied.[13] Coverage was capped at 95% or higher if a country has ever achieved greater than 95% in MCV1 or MCV2 coverage. Uptake is a variable that estimates how quickly a country is able to introduce a routine vaccine into its national immunization system. Uptake of MCV1 and MCV2 vaccine was not considered a relevant variable as all countries were forecasted to introduce prior to 2030. [5] The coverage rate and growth were calculated on a yearly basis. A 100% coverage for SIAs was applied to estimate the PDR for SIA; such an assumption was also based on MI4A methodology, which considers the standard practice to procure SIA doses for the entire population.[13] SIAs were assumed to be implemented in one single year (e.g., uptake at 100%) with the exception of countries that have historically implemented SIAs over 2-3 years, including Democratic Republic of Congo, Egypt, Ethiopia, Indonesia, Nigeria, Pakistan, and Philippines. This is in line with the MI4A methodology.[13]

#### Forecast the frequency of SIAs

##### Data: annex.xls, sheets 8a-8b

The frequency of the SIAs targeting 9–59-month-old children were forecasted until 2040 based on the forecasted MCV2 coverage using MI4A methodology: (i) if countries had a MCV2 coverage of <60%, they were projected to conduct an SIA every 2 years; (ii) if countries had an MCV2 coverage between 60-80%, they were assumed conducting an SIA every 3 years; and (iii) if countries had a MCV2 coverage of > 80%, SIA were foreseen every 4-5 years.[13] Countries were forecasted to stop conducting SIAs when their MCV2 coverage reached 90% for 3 consecutive years.[4]

#### Calculate buffer stock and wastage

##### Data: annex.xls, sheet 5

WHO guidance was followed to estimate buffer stock and wastage rates for the N/S MR vaccine.[16, 17] Annual buffer stock was calculated as 25% of the difference in demand from the current and the prior year of vaccine routine use, negative values were transformed to zero. The following wastage rates were applied for N/S vaccines to be delivered routinely in infants: 1-dose vial 5%, 5-dose vial 15%, and 10-dose vial 40%. The following wastage rates were applied for N/S vaccines to be delivered in SIAs: 1-dose vial 1%, 5-dose vial 10%, 10-dose vial 10%. [16, 17] The wastage rate for MR-MAPs was set at 1% due to their assumed increased thermostability, single dose presentation, and shelf life.[16, 17]

Figure S1 provides an overview of the methodology and assumptions to extrapolate MCV routine immunization doses to 2040.

**Figure S1:**
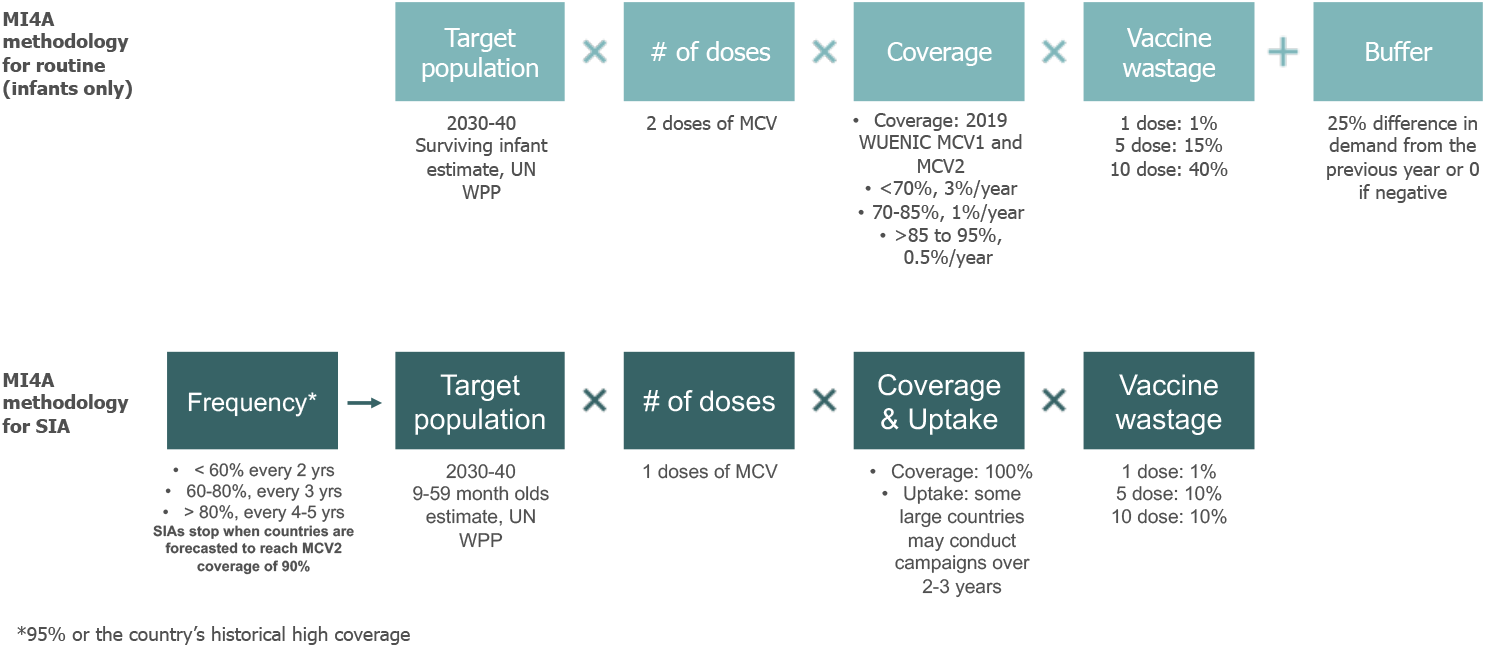
Methodology and assumptions to estimate MCV routine immunization and SIA PDR for 2030-2040 (Step 1). MR: measles and rubella; SIA: supplementary immunization activities; PDR: programmatic doses required; MI4A: Market Information for Access to Vaccines; UN: United Nations; WPP: World Population Prospectus; WUENIC: WHO/UNICEF Estimates of National Immunization Coverage; MCV1: 1^st^ dose of a measles containing vaccine; MCV2: 2^nd^ dose of a measles containing vaccine.

### Step 2: Estimate MR-MAP PDR

#### Create country archetypes

##### Data: annex.xls, sheet 1a

The MR-MAP demand forecast utilized a hybrid method where assumptions were developed for four country archetypes, and individually, for 16 key countries. These countries included the ten countries with the largest populations, the ten countries with the largest number of unimmunized children based on MCV1 coverage, and six countries that are judged high priority by the Measles and Rubella Initiative (M&RI) and/or Gavi, the Vaccine Alliance (Gavi). Using these criteria, 16 key countries were identified, and these countries account for approximately 50% of the under 5 year old population.[18] (Figure S2).

**Figure S2:**
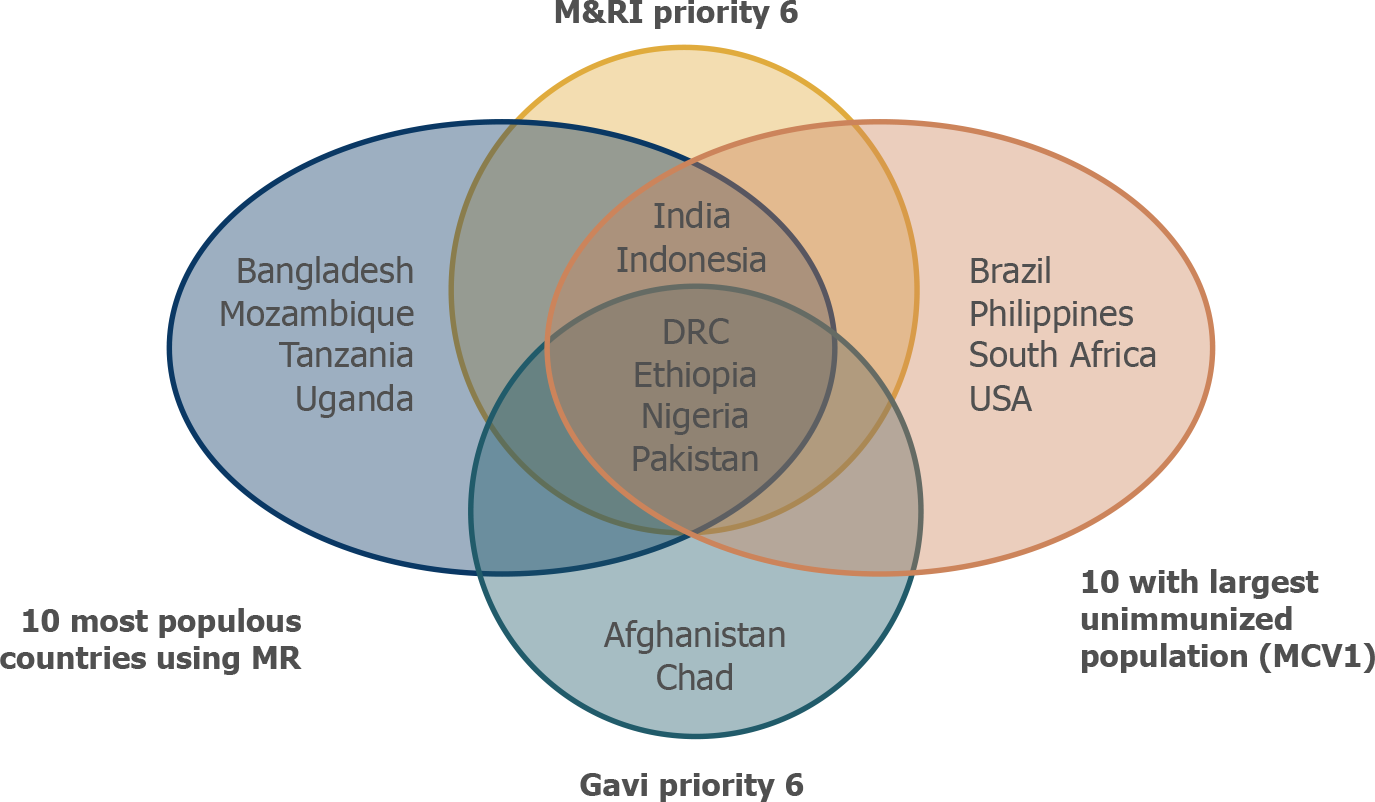
Identification of 16 key countries. M&RI: Measles And Rubella Initiative; MR: measles and rubella; DRC: Democratic Republic of the Congo; USA: United States of America; MCV1: 1^st^ dose of a measles containing vaccine.

The remaining 167 countries were grouped based on their forecasted MCV use and their WHO regions to reflect potential differences in health systems and how vaccines are delivered. This resulted in four country archetypes:

A. Countries that exclusively use MMR / MMRV

B. Countries that use MMR / MMRV in routine immunization but may use MR / M for SIAs

C. Countries that use M or MR in WHO’s African & Eastern Mediterranean Region

D. Countries that use M or MR in WHO’s Southeast Asian & Western Pacific Region

Where country assumptions were unavailable, the weighted average estimated for the group which included that country was applied.

#### Estimate country adoption year of MR-MAPs

##### Data: annex.xls, sheet 8d

To estimate the proportion of global MCV demand delivered by MAPs, a framework to predict the year of country adoption of MR-MAPs was developed, based on the following three parameters: (i) the year of introduction of MCV2, rubella, pneumococcal conjugate, rotavirus, and human papillomavirus vaccines; (ii) the total disease burden of measles and rubella (2019 reported annual cases and deaths of measles, 2019 measles incidence rate, and 2010 number of estimated rubella cases), and (iii) the percentage of total expenditure on vaccines funded by the government (%) for the last available year or the forecasted Gavi eligibility status in 408 2030. [19-23]

The predictive framework considers the variables and assigns points distributed by quartiles using the data for each country. For example, the introduction of rotavirus vaccines occurred between 2006 to the present day; thus, those countries that introduced the vaccine within the first quartile or 25^th^ percentile of all years 2006-2022, i.e. years 2006 to 2011, were assigned 4 points, second quartile (i.e., years 2011 to 2014) were assigned 3 points, third quartile (i.e., years 2014-2017) were assigned 2 points, and fourth quartile (i.e., 2017-present day) were assigned 1 point. The quartiles were calculated for each variable with the points assigned in a similar fashion. Then, the scores for each variable were totalled by country, and based on this score, the countries were assigned an adoption year which was equally spread across the 11-year time period of 2030-40. This resulted in ∼16 countries adopting MR-MAPs each year. This framework predicts that countries would adopt MR-MAPs earlier if they had a history of early introduction of new vaccines, had a high level of measles and rubella burden, and had sufficient financial resources devoted to immunization or were anticipated to receive donor support. The variables were considered equally for the base estimates.

#### Market penetration of MR-MAPs

To estimate the proportion of global MCV demand delivered by MAPs, an MR-MAP market penetration rate, varied by country groups, was applied to the total MCV PDR calculated from Step 1. A market penetration of 5% was assumed for Group A as these countries would largely continue using MMR and MMRV for the majority of their programmes, but potentially utilize MR-MAPs to vaccinate special and vulnerable populations, such as migrants. A market penetration of 30% was used for Group B countries based on the historical use of MR vaccines in MMR/MMRV countries, which estimated that MR N/S accounted for approximately 30% of total MCV demand for these countries.[24] Finally, a market penetration of 80% was applied for Groups C and D based on the assumption that these countries would unlikely switch fully to MR-MAPs.[24] In the absence of available data, the market penetration of MR-MAPs was discussed and agreed upon by a group experts and consulted with country immunization experts.

Figure S3 provides an overview of the methodology and assumptions for Step 2.

**Figure S3:**
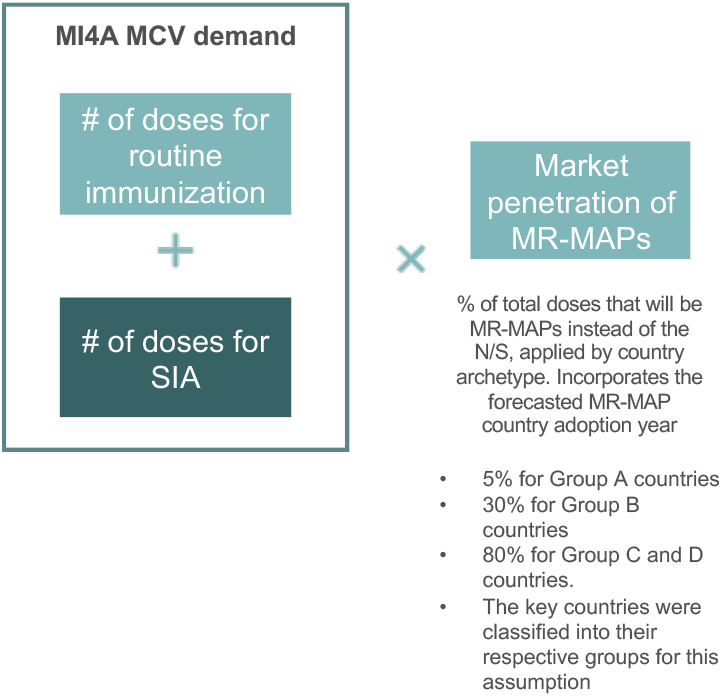
Step 3 Split global MCV PDR by presentation and vaccine type. SIA: supplementary immunization activities; MI4A: Market Information for Access to Vaccines; MR: measles rubella; MR-MAP: measles and rubella microarray patches; N/S: needle and syringe.

### Step 3: Estimate additional PDR due to increased reach of MR-MAPs Characterise the considered target population and immunization strategies

#### Data: annex.xls, sheets 3a-3e

To estimate the full PDR of MR-MAPs, we calculated the size of additional populations that could be reached by MR-MAPs, including wastage and buffer stock. These populations included those who are not receiving MCV vaccines due to missed opportunities for vaccinations (MOV) or living in hard-to-reach areas. We used data from published and unpublished data sources, such as country comprehensive multi-year plans, EPI reviews, Gavi joint appraisal reports, World Bank estimates for population living in urban slums and rural areas, and 2019 United Nations High Commissioner for Refugees estimates for population of concern.[18, 25-30]

To estimate the size of the hard-to-reach populations, the children between 0-2, and 2-15 years old, who lived in urban slums, remote rural, and security compromised settings were included. The relevant data was obtained using the above data sources and then stratified by age based on United Nations World Population Prospectus.[18, 25, 26, 28, 29] The estimates for hard-to-reach populations were held constant for the entire forecasting period given the high level of uncertainty in how these estimates may evolve over the next 20 years.

An MOV refers to any contact with health services by a child or adult who is eligible for vaccination, which does not result in the person receiving one or more of the vaccine doses for which he or she is eligible. The estimated size of the MOV population was calculated as 2% of children under 2 years of age per United Nations World Population Prospectus from 2030-40. This percentage was derived from published MOV literature and validated by expert opinion which considered the timeliness of vaccination and methodology to develop WUENIC estimates to avoid overestimating the MOV population.[18, 28, 29, 31] The MOV percentage was held constant for the forecasting period.

The following immunization strategies were used to reach the MOV and hard-to-reach populations: (i) two doses delivered as part of additional routine immunization for children less than two years of age in MOV and hard-to-reach populations (e.g., security compromised, remote rural, and urban slum populations); and (ii) two doses delivered through SIAs in children between the ages of two to 15 years in hard-to-reach populations.[18, 25-29]

### Forecast coverage of hard-to-reach and MOV populations

#### Data: annex.xls, sheets 4c-4d

For the hard-to-reach and MOV populations, the coverage was estimated based on the expert opinion of members of the Working Group of Experts on MR-MAPs and MI4A Advisory Group. While MR-MAPs were assumed to increase the MR reach in hard-to-reach and MOV populations, their ability to fully address all programmatic barriers was considered unlikely. Hence, for the hard-to-reach and MOV populations, a coverage of 20% was applied in a routine immunization setting for both doses, and 10% was applied for the SIAs. These estimates are conservative given the lack of information on the MAP reach of hard-to-reach and MOV populations. These estimates were also held constant for the forecasting period given the lack of data available to inform this assumption. Uptake of MR-MAPs was assumed to be 100% for both routine and the one-time catch-up campaign of MOV and hard-to-reach populations.

Figure S4 provides an overview of the methodology and assumptions to calculate the additional PDR due to the use of MAPs.

**Figure S4:**
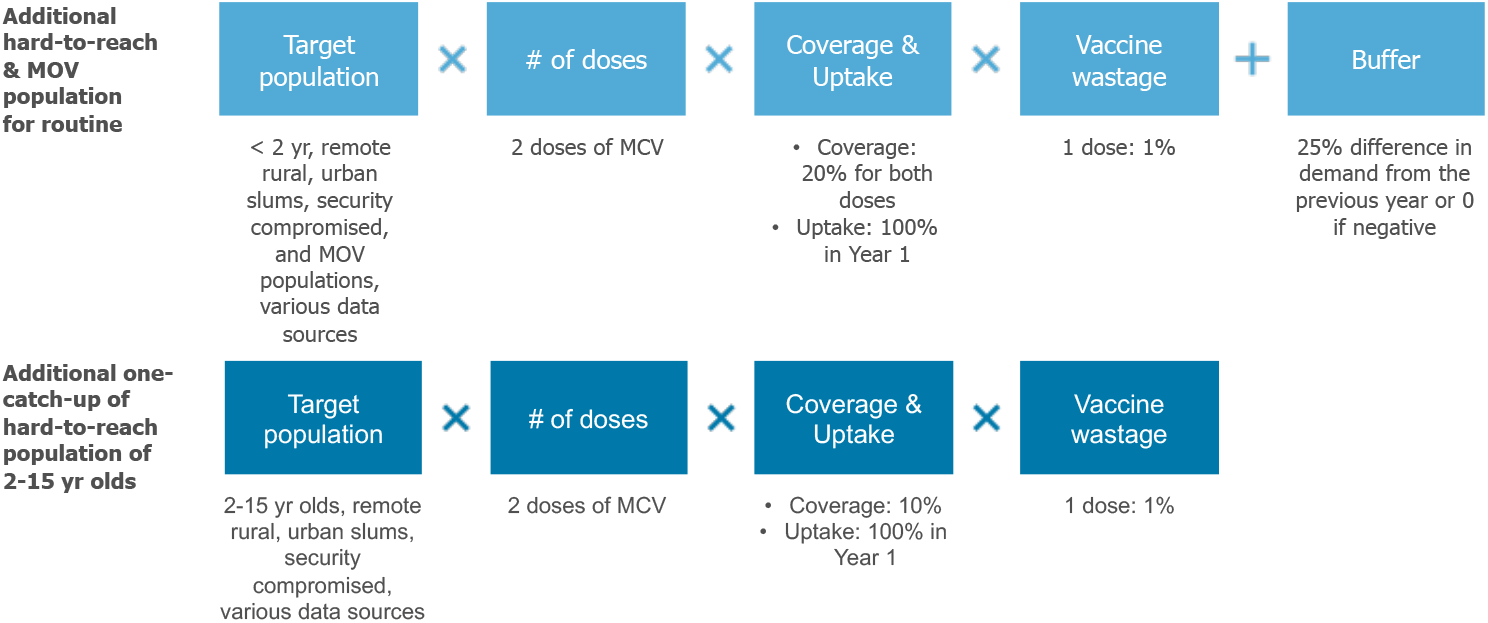
Methodology and assumptions utilized to calculate additional reach of MR-MAPs in the hard-to-reach and MOV population. MR-MAPs: measles and rubella microarray patches; MOV: missed opportunities for vaccination; MCV: measles containing vaccines; yr: years.

### Step 4: Estimate the proportion of MR-MAP PDR delivered by Use Cases

#### Data: annex.xls, sheets 6a-6c

To estimate where MR-MAPs would be delivered (fixed post with full cold chain capabilities versus limited or no cold chain capabilities) and by whom (health workers, community health workers), we have applied the previously developed Use Cases 1-4 (text box 2) to the MR-MAP PDR developed in step 3. We applied assumptions about the proportion of vaccines delivered in the mentioned locations and by the mentioned personnel. The assumptions relating to the number of nurses or midwives and community health workers were obtained from the Global Health Workforce statistics and supported by unpublished literature, such as country comprehensive multi-year plans. The assumptions related to vaccine delivery in fixed posts or during outreach were largely obtained from unpublished literature such as from country comprehensive multi-year plans, Expanded Programme on Immunization (EPI) reviews, Gavi joint appraisal reports, validated during interviews with country EPI managers and immunization focal points. [32] The data was not adjusted for the period 2030-2040, nor it was changed to reflect the introduction of MAPs. Figure S5 provides an overview of the methodology and assumptions.

**Figure S5:**
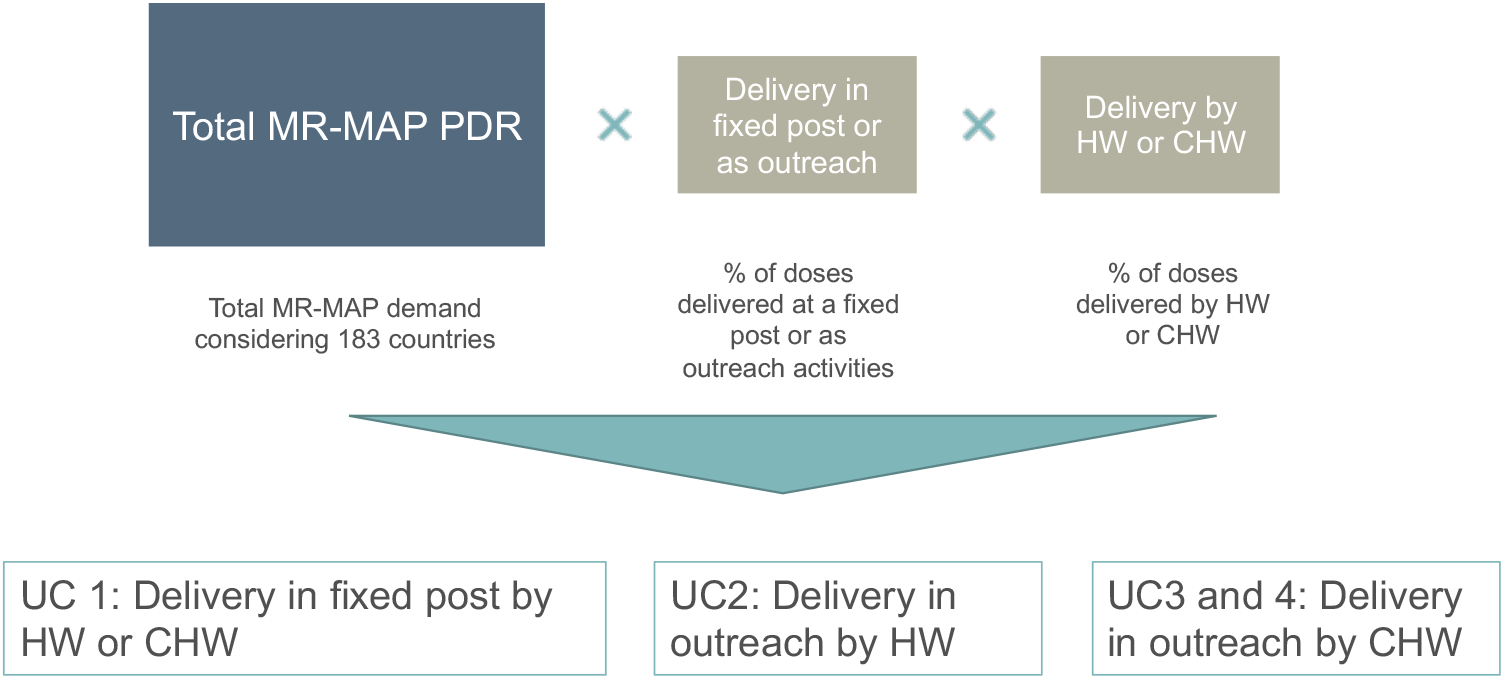
Splitting MR-MAP PDR by Use Case. MR-MAP: measles and rubella microarray patches; PDR: programmatic doses required; HW: health workers; CHW: community health workers; UC: use case.

### Step 5: Explore uncertainty of MR-MAP PDR with scenarios

#### Data: annex.xls, sheet 8e

Given the high level of uncertainty in how global PDR will evolve for MR-MAPs between 2030-2040, three scenarios were explored (text box 4).

##### Text box 4: Description of the scenarios

- Scenario 1: **Base**: utilizes 5-step demand forecasting methodology described above.
- Scenario 2: **Regional MR-MAP pilots**: Five countries in each WHO region conduct sub-national pilots with MR-MAPs for 2 years prior to national implementation. All other countries do not adopt MR-MAPs until the regional pilots are completed.
- Scenario 3: **Accelerated adoption in countries with the greatest need**: Countries with the lowest MCV1 coverage and highest disease burden for measles and rubella are forecasted to adopt MR-MAPs earlier.

Note that there were no constraints applied to any of the scenarios.

The scenarios were developed based on the input of the Working Group of Experts on MR-MAPs and MI4A Advisory Group. Scenario 1 serves as the **base scenario** and utilizes the forecasting methodology described above to estimate PDR and is described in Steps 1-4. Scenario 2, **regional MR-MAP pilots**, considered if MR-MAPs would be rolled out as ‘sub-national pilots’ in the first five countries in each WHO region targeting only 5% of the surviving infant population for the first two years. These countries would introduce MR-MAPs nationally in year 3 while all other countries would delay the introduction of MR-MAPs until the regional pilots were completed (5 years from the beginning of the pilot). Scenario 3 explored if MR-MAPs were adopted first in countries that had the **greatest need** based on their MCV1 coverage and their disease burden of measles and rubella. This was achieved by utilizing the same predictive framework to forecast adoption year, but instead of the variables being equally weighted, the variables for MCV1 coverage and measles and rubella burden were given twice the weight compared to the other variables. Based on the revised weights, a new adoption year was calculated.

## Supplemental Annex B: Methodology and results of sensitivity analyses

Sensitivity analyses were conducted on specific variables where no or limited data existed or if there were concerns regarding the quality of the data highlighted by the Expert group on MR-MAPs and the MI4A Advisory Group. Four types of sensitivity analyses were conducted on (i) additional hard-to-reach and MOV populations, (ii) proportion of additional hard-to-reach and MOV populations reached by MR-MAPs, (iii) delivery location, and (iv) service provider.

These analyses aimed to identify which variables have the highest impact on the demand forecast so that efforts could be focused on filling the most impactful evidence gaps to improve the accuracy and reliability of the demand forecast.

For simplicity, the variables were considered independently and explored using a plus or minus 20% change. 20% was chosen as the MI4A MCV analysis shows an annual change in total MCV routine and SIA PDR of ∼22-28%.[13]

The first sensitivity analysis conducted reviewed the estimated hard-to-reach and MOV populations. Based on expert feedback, there were concerns that the definition of these hard-to-reach and MOV populations may either over or underestimate the population and/or there may be overlap between the different populations. Figure S6 provides an overview of the results of the sensitivity analyses for these populations.

**Figure S6:**
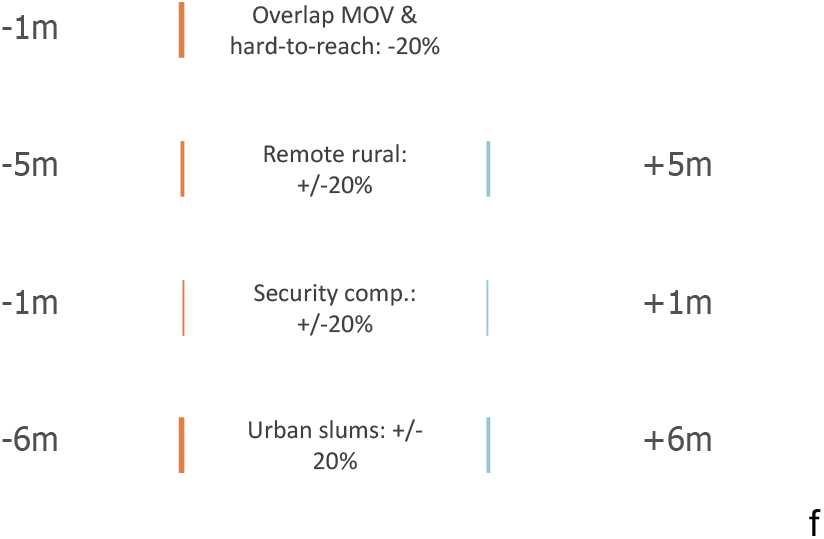
Overview results of sensitivity analyses on hard-to-reach and MOV populations. MOV: missed opportunities for vaccination.

The estimated hard-to-reach and MOV populations could have an impact of 1 to 6 million PDR on the overall demand forecast. As this accounts for less than 2% of the total annual PDR once a steady state is reached, it is unlikely that inaccuracies in these population estimates will have significant impact on the overall demand forecast.

Next, we reviewed the proportion of the hard-to-reach and MOV populations that could be reached by a MR-MAP but not a N/S presentation for routine immunization and a one-time catch-up campaign. In contrast, the coverage did have a significant impact on the estimated PDR impacting demand by -84 million doses or by +207 million doses. This highlights the importance in understanding whether MR-MAPs, given their innovative product characteristics, can actually close immunization gaps and reach the zero-dose or under-vaccinated populations. Figure S7 provides the results of this sensitivity analysis.

**Figure S7:**
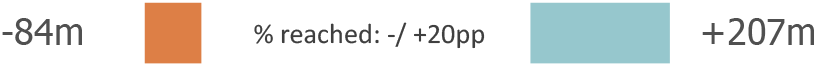
Results of the +/-20% sensitivity analysis on the proportion of hard-to-reach and MOV populations that could be reached by a MR-MAP but not a N/S. MOV: missed opportunities for vaccination; MR-MAP: measles and rubella microarray patches; N/S: needle and syringe; pp: percentage points; m: millions.

Next, we looked at how percentage changes impacted the UC dimensions of delivery location and service provider. While this sensitivity did not change the total estimated PDR for MR-MAPs, it did change how countries would be utilizing the MR-MAPs and the proportion of doses delivered in different UCs.

An increase in the delivery at fixed post by 20% resulted in an increase in UC1 (+41%) with decrease of UC2 and UC3+4 (−38% and -37%, respectively), which focus more on outreach activities. In contrast a decrease of 10% delivery in fixed post resulted in a decrease in UC1 (−21%) and increase in UC2 and UC3+4 (+19% for both). The delivery location implies a certain level of stability with the PDR and shows that there is interest for MR-MAPs to be delivered in various locations not only in certain settings.

Finally, considering a +/-20 percent change in whether community health worker could deliver MR-MAPs impacted the PDR between UC2 and UC3+4. If 20% more community health workers are able to deliver MR-MAPs this could create an increase in UC3+4 of ∼10% and a decrease in UC2 of -7%. In comparison, if community health workers are unable to deliver MR-MAPs, then this may place more importance on UC2 (+8%) compared to UC3+4 (−12%). Country consultations have indicated that there could be legal and technical hurdles to allowing community health workers to deliver MR-MAPs. These hurdles may reduce a MR-MAP’s value proposition to expand the potential vaccination workforce to reach the zero-dose and under-immunized as it will not allow for differentiation from the N/S presentation. Given that these sensitivity analyses show ∼10% change in the UCs, it highlights the importance of understanding where MR-MAPs will be used and who can deliver MR-MAPs as this could impact the anticipated value of a MR-MAP ultimately affecting investment decisions. Figure S8 provides an overview of the results of the sensitivity analyses on the UC dimensions.

**Figure S8:**
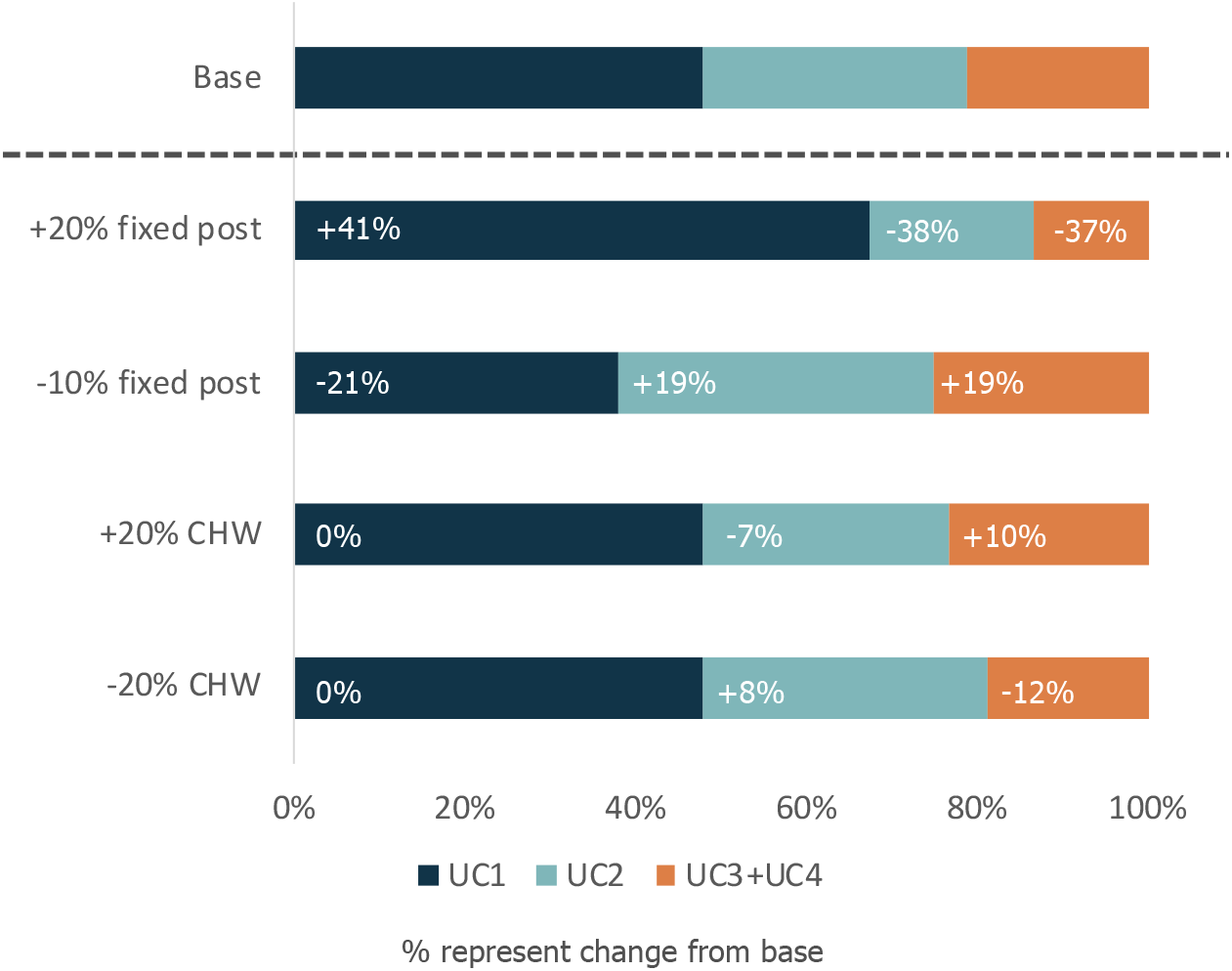
Results of the +/- 20% sensitivity analysis on UC dimensions of delivery location and service provider. UC: use case; CHW: community health worker.

## Supplemental Annex C: Summary tables of the demand forecasting results

**Table S1:**
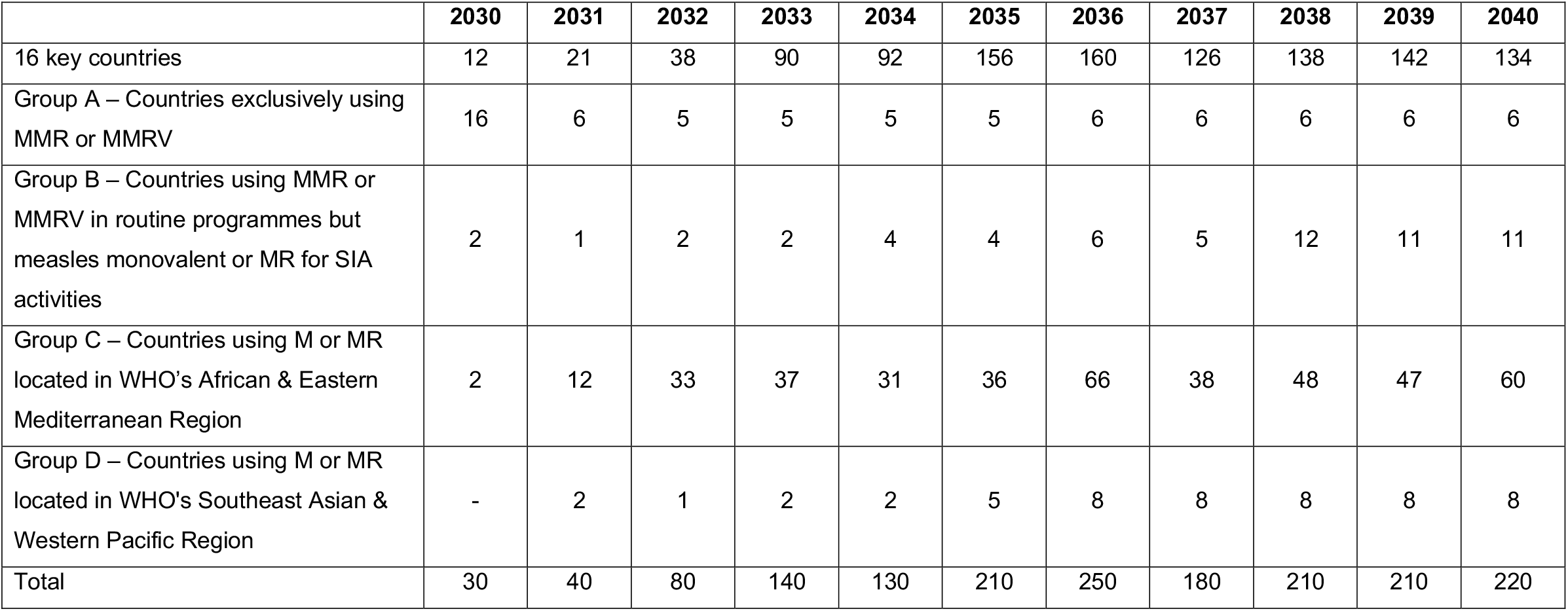
Estimated programmatic doses required of MR-MAPs by country archetype and year in millions. M: measles; MR: measles, 613 rubella; MMR: measles, mumps rubella; MMRV: measles, mumps, rubella, varicella; SIA: supplementary immunization activities; WHO: World 614 Health Organization.

**Table S2:**
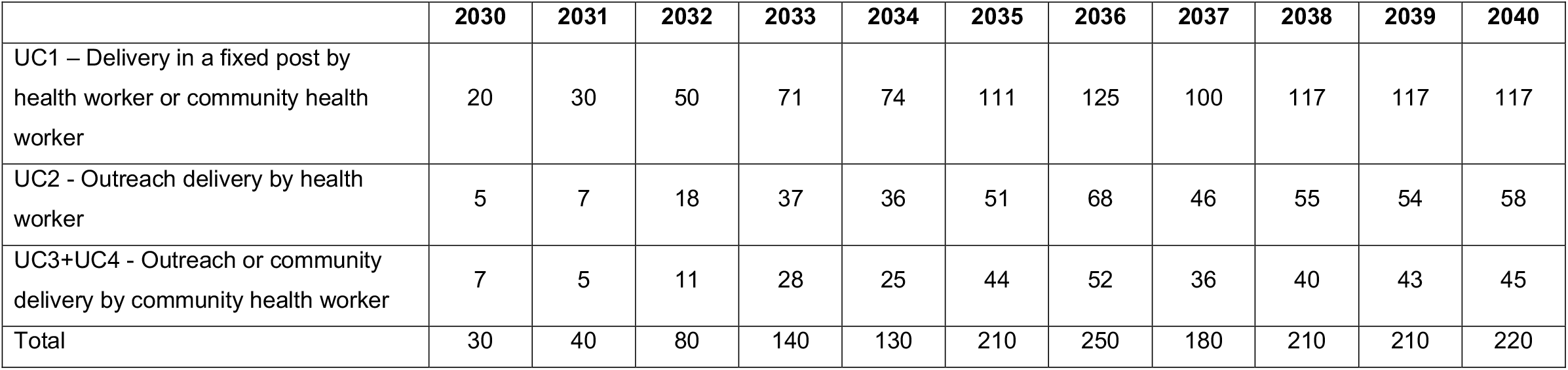
Estimated programmatic doses required of MR-MAPs by use cases and year in millions. UC: use case.

**Table S3:**
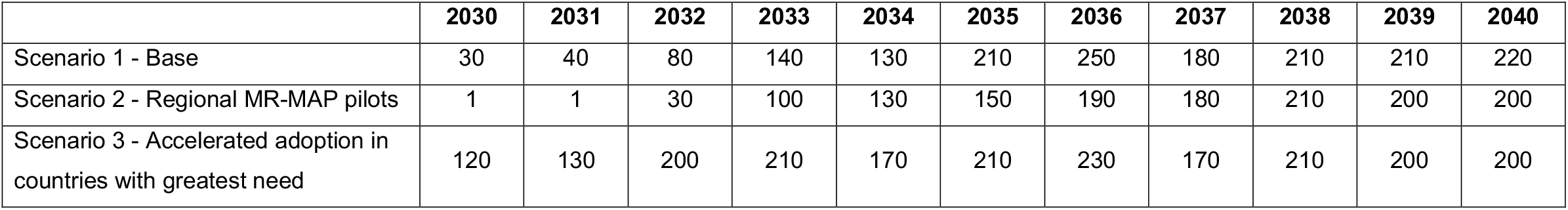
Estimated programmatic doses required of MR-MAPs by scenario and year in millions. MR-MAP: measles and rubella microarray patches.

## Notes

**Conflict of interest:** all authors declare no conflict of interest

### Competing Interest Statement

The authors have declared no competing interest.

