## Supplemental Annex for "Estimating the future global dose demand for Measles-Rubella microarray patches"

Figure 6 provides an overview of the methodology and assumptions to extrapolate MCV routine immunization doses to 2040.

**Figure 6: Methodology and assumptions to estimate MCV routine immunization and SIA PDR for 2030-2040 (Step 1)**

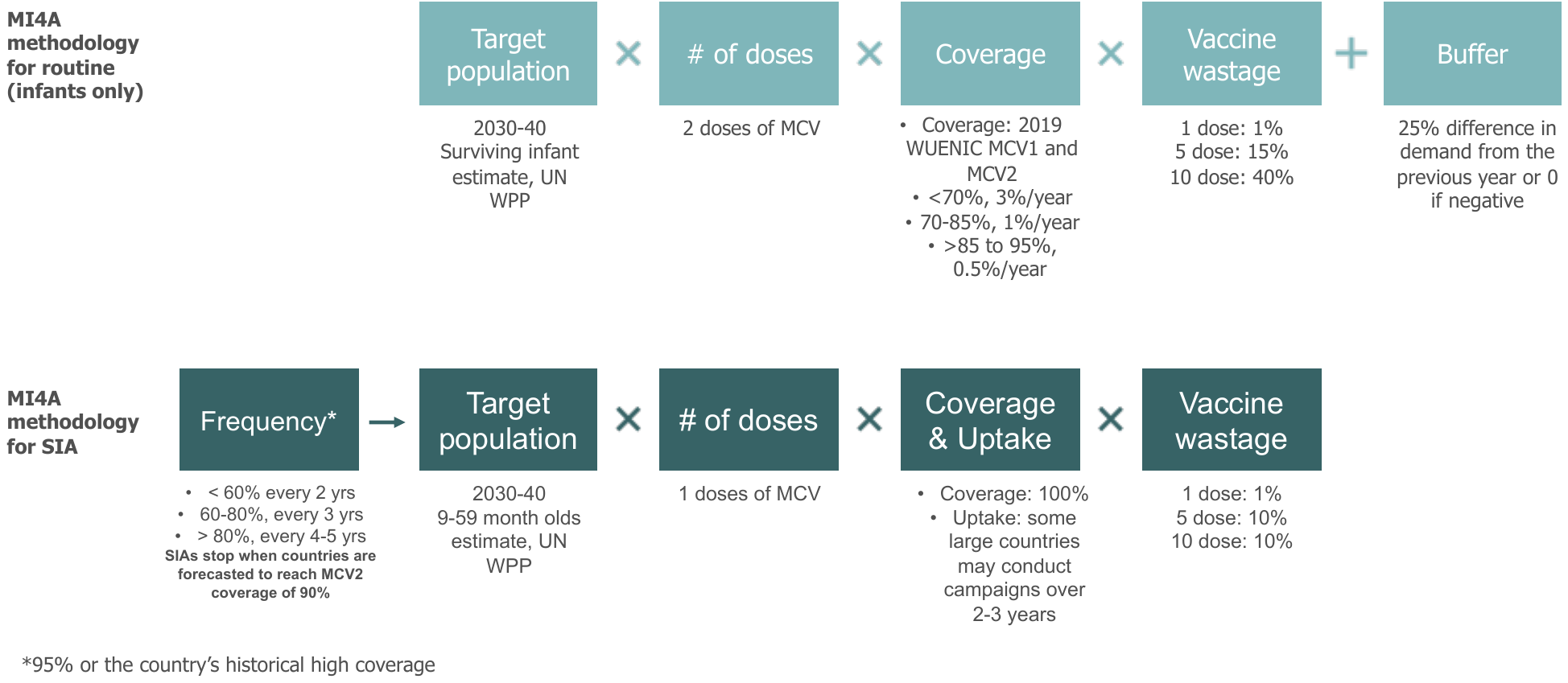

#### Step 2: Estimate MR-MAP PDR

#### D. Create country archetypes

The MR-MAP demand forecast utilized a hybrid method where assumptions were developed for four country archetypes, and individually, for 16 key countries. These countries included the ten countries with the largest populations, the ten countries with the largest number of unimmunized children based on MCV1 coverage, and six countries that are judged high priority by the Measles and Rubella Initiative (M&RI) and/or Gavi, the Vaccine Alliance (Gavi). Using these criteria, 16 key countries were identified, and these countries account for approximately 50% of the under 5 year old population.[8] (Figure 7).

**Figure 7 Identification of 16 key countries**
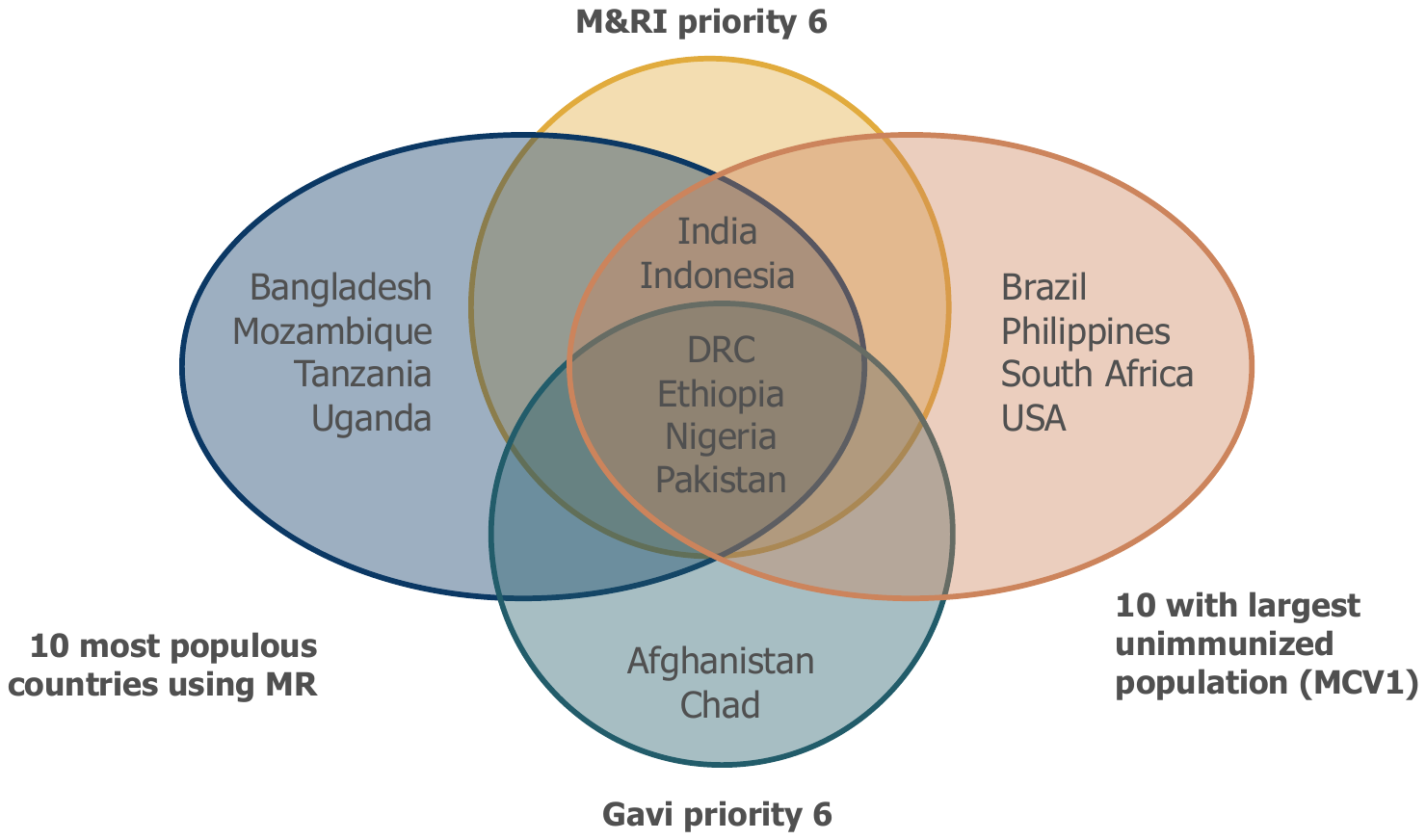

The remaining 167 countries were grouped based on their forecasted MCV use and their WHO regions to reflect potential differences in health systems and how vaccines are delivered. This resulted in four country archetypes:

A. Countries that exclusively use MMR / MMRV
B. Countries that use MMR / MMRV in routine immunization but may use MR / M for SIAs
C. Countries that use M or MR in WHO’s African & Eastern Mediterranean Region
D. Countries that use M or MR in WHO's Southeast Asian & Western Pacific Region

Where country assumptions were unavailable, the weighted average estimated for the group which included that country was applied. See supplemental annex.xls, sheet 1 to see what archetype was assigned to each country.

#### E. Estimate country adoption year of MR-MAPs

To estimate the proportion of global MCV demand delivered by MAPs, a framework to predict the year of country adoption of MR-MAPs was developed, based on the following three parameters: (i) the year of introduction of MCV2, rubella, pneumococcal conjugate, rotavirus, and human papillomavirus vaccines (supplemental annex.xls, sheet 8a); (ii) the total disease burden of measles and rubella (2019 reported annual cases and deaths of measles, 2019 measles incidence rate, and 2010 number of estimated rubella cases, (supplemental annex.xls, sheet 8b)), and (iii) the percentage of total expenditure on vaccines funded by the government (%) for the last available year or the forecasted Gavi eligibility status in 2030 (supplemental annex.xls, sheet 8c).[9-13]

**Figure 8: Step 3 Split global MCV PDR by presentation and vaccine type**

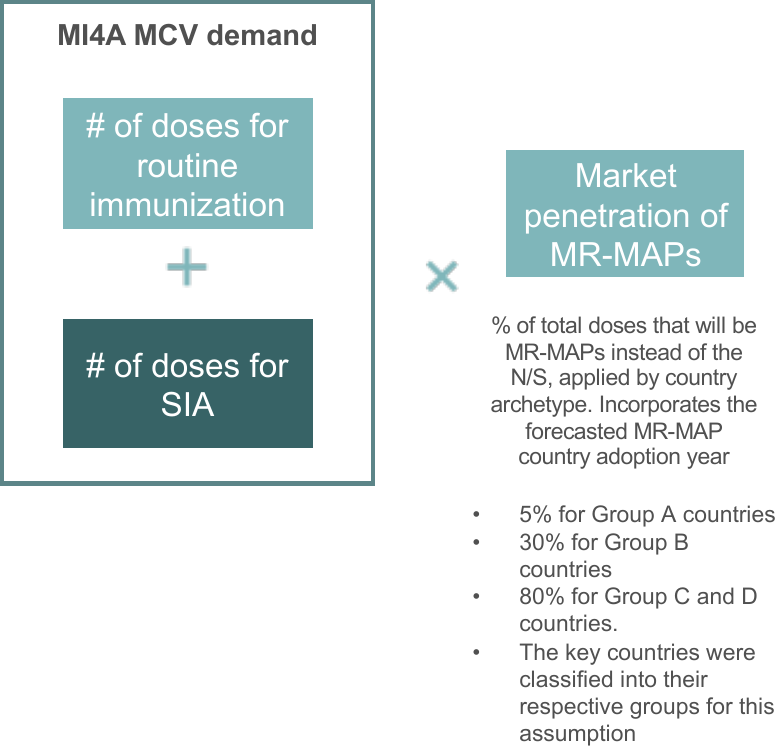

#### Step 3: Estimate additional PDR due to increased reach of MR-MAPs

#### A. Characterise the considered target population and immunization strategies

Figure 9 provides an overview of the methodology and assumptions to calculate the additional PDR due to the use of MAPs.

**Figure 9: Methodology and assumptions utilized to calculate additional reach of MR-MAPs in the hard-to-reach and MOV population.**

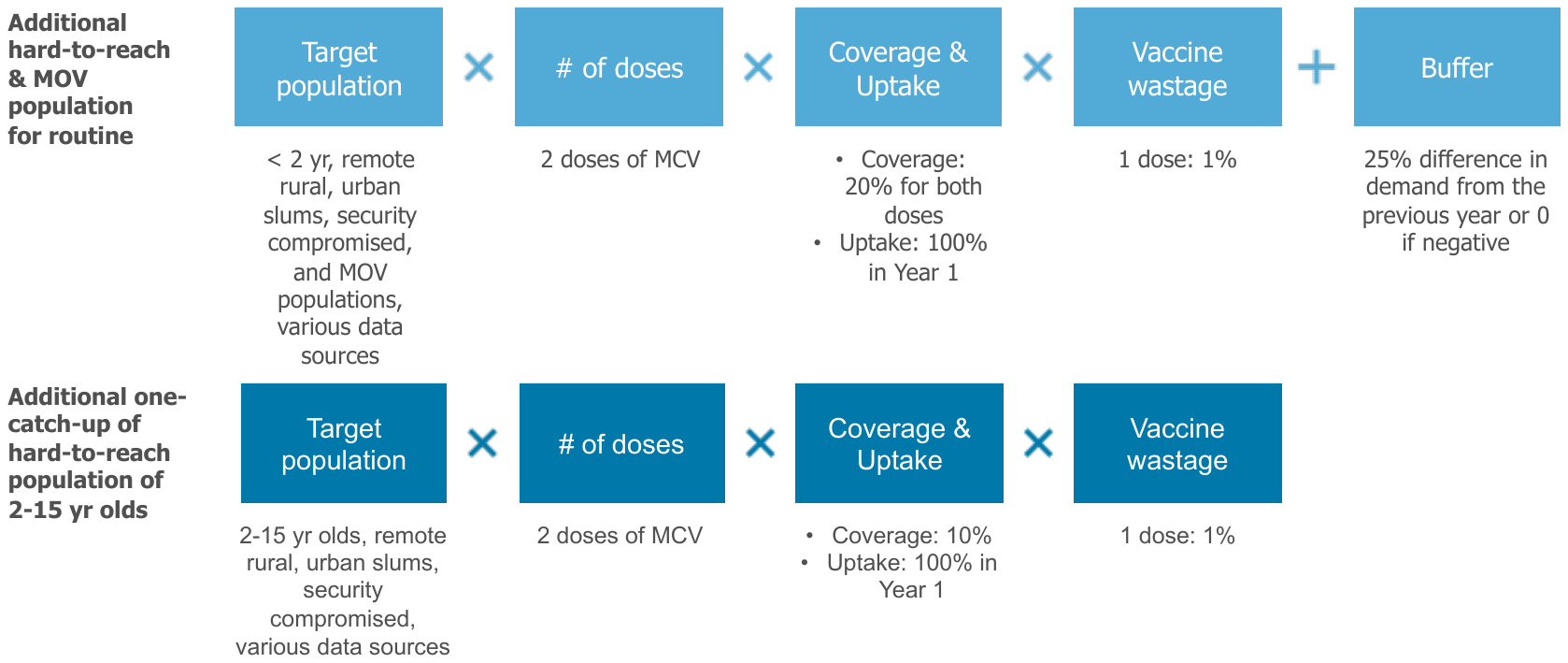

##### Step 4: Estimate the proportion of MR-MAP PDR delivered by Use Cases

To estimate where MR-MAPs would be delivered (fixed post with full cold chain capabilities versus limited or no cold chain capabilities) and by whom (health workers, community health workers), we have applied the previously developed Use Cases 1-4 (see Box 1) to the MR-MAP PDR developed in step 3. We applied assumptions about the proportion of vaccines delivered in the mentioned locations and by the mentioned personnel. The assumptions relating to the number of nurses or midwives and community health workers were obtained from the Global Health Workforce statistics and supported by unpublished literature, such as country comprehensive multi-year plans (supplemental annex.xls, sheet 6c). The assumptions related to vaccine delivery in fixed posts or during outreach were largely obtained from unpublished literature such as from country comprehensive multi-year plans, Expanded Programme on Immunization (EPI) reviews, Gavi joint appraisal reports, validated during interviews with country EPI managers and immunization focal points (supplemental annex.xls, sheet 6a-6b). [22] The data was not adjusted for the period 2030-2040, nor it was changed to reflect the introduction of MAPs.

Figure 10 provides an overview of the methodology and assumptions.

**Figure 10: Splitting MR-MAP PDR by Use Case**

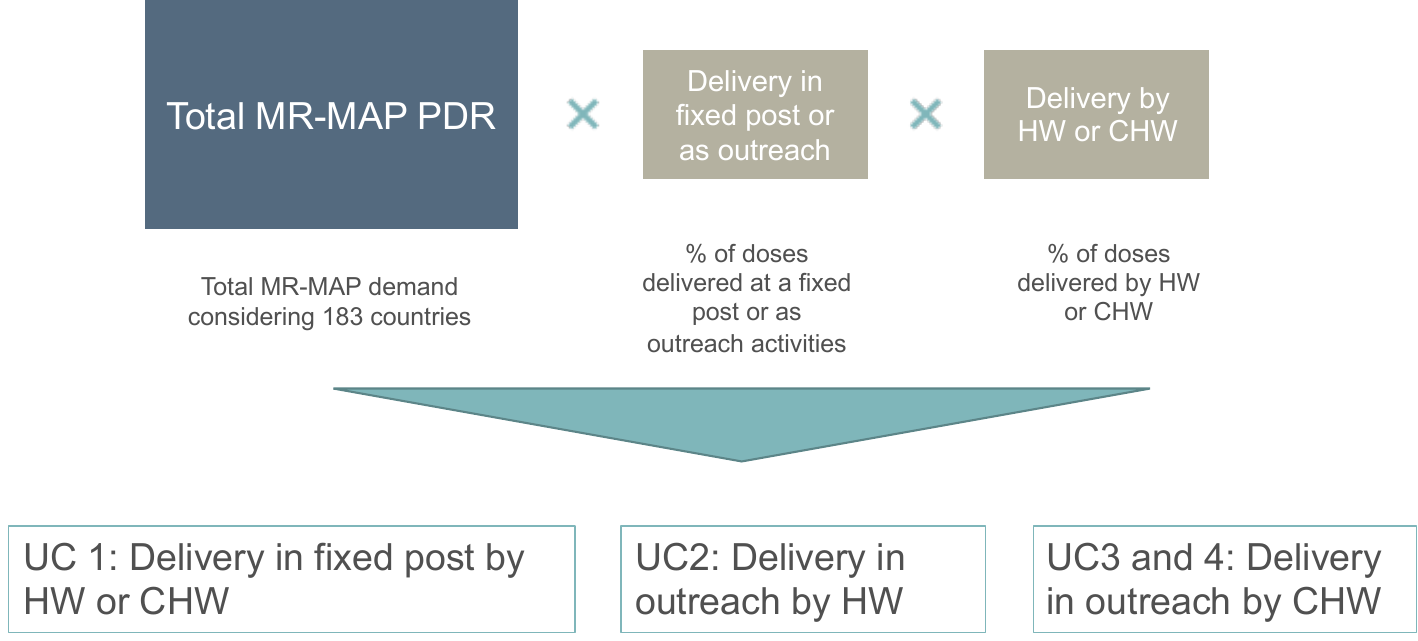

#### Step 5: Explore uncertainty of MR-MAP PDR through the use of scenarios

f

**Text box 2: Description of the scenarios**

**Figure 11: Overview results of sensitivity analyses on hard-to-reach and MOV populations**

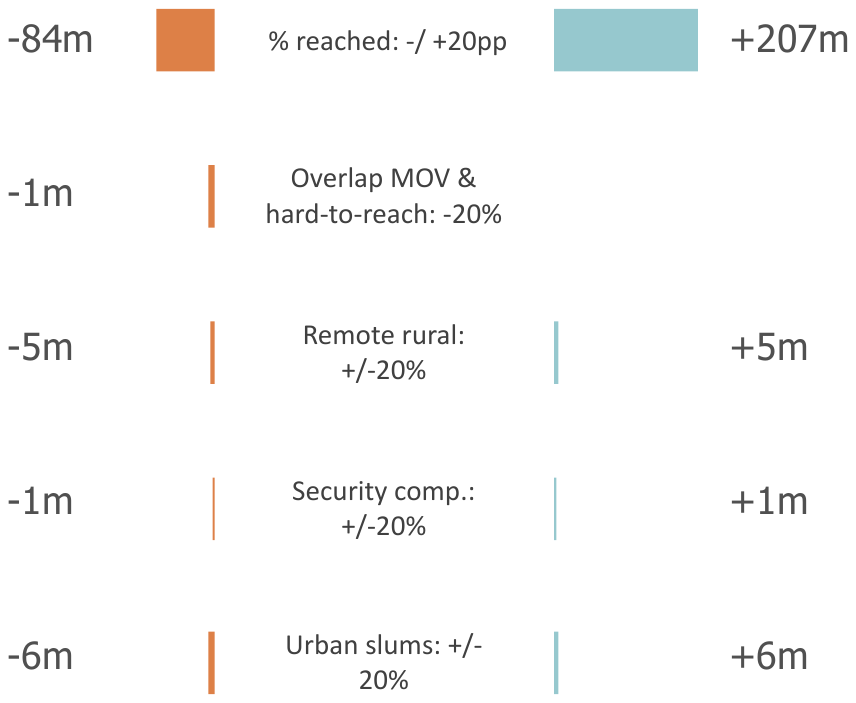

The estimated hard-to-reach and MOV populations could have an impact of 1 to 6 million PDR on the overall demand forecast. As this accounts for less than 2% of the total annual PDR once a steady state is reached, it is unlikely that inaccuracies in these population estimates will have significant impact on the overall demand forecast.

Next, we reviewed the proportion of the hard-to-reach and MOV populations that could be reached by a MR-MAP but not a N/S presentation for routine immunization and a one-time catch-up campaign. In contrast, the coverage did have a significant impact on the estimated PDR impacting demand by -84 million doses or by +207 million doses. This highlights the importance in understanding whether MR-MAPs, given their innovative product characteristics, can actually close immunization gaps and reach the zero-dose or under-vaccinated populations. Figure 12 provides the results of this sensitivity analysis.

**Figure 12: Results of the +/-20% sensitivity analysis on the proportion of hard-to-reach and MOV populations that could be reached by a MR-MAP but not a N/S**

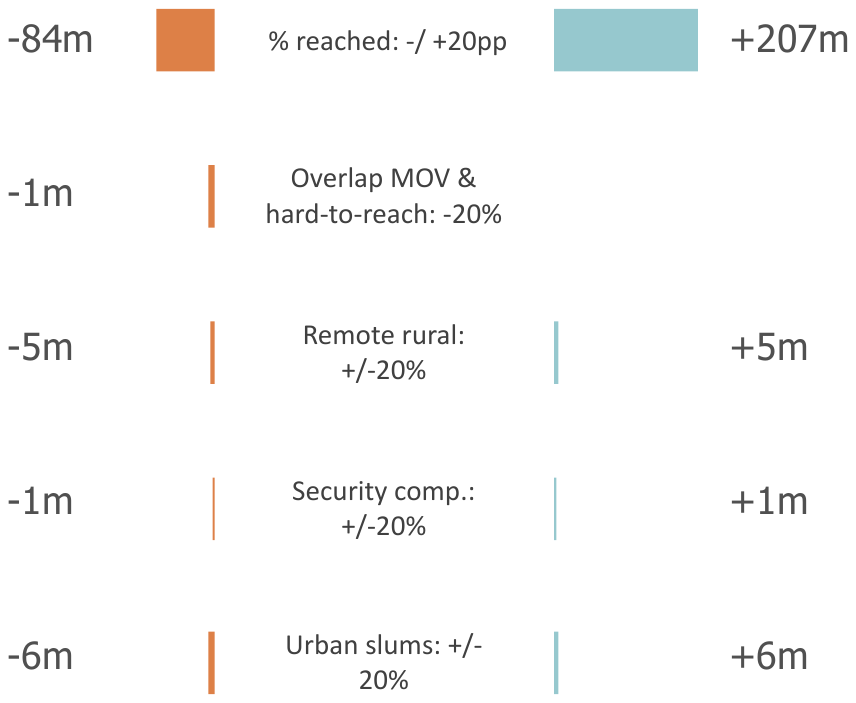

Next, we looked at how percentage changes impacted the UC dimensions of delivery location and service provider. While this sensitivity did not change the total estimated PDR for MR-MAPs, it did change how countries would be utilizing the MR-MAPs and the proportion of doses delivered in different UCs.

An increase in the delivery at fixed post by 20% resulted in an increase in UC1 (+41%) with decrease of UC2 and UC3+4 (-38% and -37%, respectively), which focus more on outreach activities. In contrast a decrease of 10% delivery in fixed post resulted in a decrease in UC1 (-21%) and increase in UC2 and UC3+4 (+19% for both). The delivery location implies a certain level of stability with the PDR and shows that there is interest for MR-MAPs to be delivered in various locations not only in certain settings.

Finally, considering a +/-20 percent change in whether community health worker could deliver MR-MAPs impacted the PDR between UC2 and UC3+4. If 20% more community health workers are able to deliver MR-MAPs this could create an increase in UC3+4 of ~10% and a decrease in UC2 of -7%. In comparison, if community health workers are unable to deliver MR-MAPs, then this may place more importance on UC2 (+8%) compared to UC3+4 (-12%). Country consultations have indicated that there could be legal and technical hurdles to allowing community health workers to deliver MR-MAPs. These hurdles may reduce a MR-MAP’s value proposition to expand the potential vaccination workforce to reach the zero-dose and under-immunized as it will not allow for differentiation from the N/S presentation.

Given that these sensitivity analyses show ~10% change in the UCs, it highlights the importance of understanding where MR-MAPs will be used and who can deliver MR-MAPs as this could impact the anticipated value of a MR-MAP ultimately affecting investment decisions. Figure 13 provides an overview of the results of the sensitivity analyses on the UC dimensions.

**Figure 13: Results of the +/- 20% sensitivity analysis on UC dimensions of delivery location and service provider**

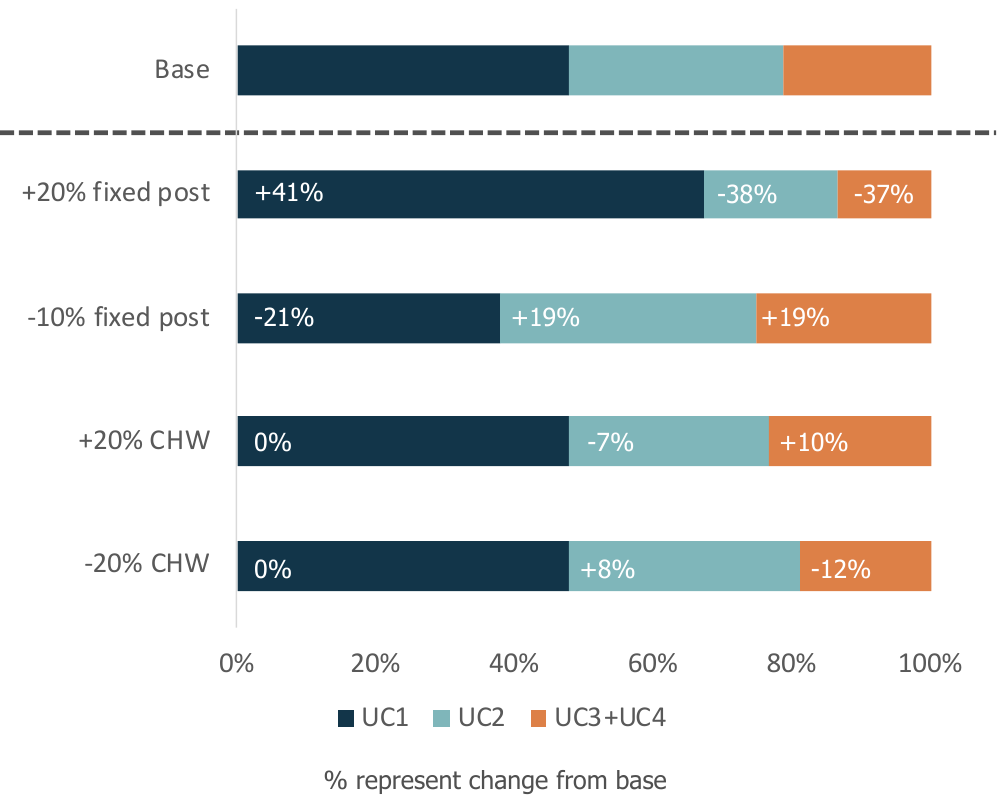

### Supplemental Annex C: Summary tables of the demand forecasting results

**Table XX: Estimated PDR by country archetype in millions**

|  | **2030** | **2031** | **2032** | **2033** | **2034** | **2035** | **2036** | **2037** | **2038** | **2039** | **2040** |
| --- | --- | --- | --- | --- | --- | --- | --- | --- | --- | --- | --- |
| 16 key countries | 12 | 21 | 38 | 90 | 92 | 156 | 160 | 126 | 138 | 142 | 134 |
| Group A – Countries exclusively using MMR or MMRV | 16 | 6 | 5 | 5 | 5 | 5 | 6 | 6 | 6 | 6 | 6 |
| Group B – Countries using MMR or MMRV in routine programmes but measles monovalent or MR for SIA activities | 2 | 1 | 2 | 2 | 4 | 4 | 6 | 5 | 12 | 11 | 11 |
| Group C – Countries using M or MR located in WHO’s African & Eastern Mediterranean Region | 2 | 12 | 33 | 37 | 31 | 36 | 66 | 38 | 48 | 47 | 60 |
| Group D – Countries using M or MR located in WHO's Southeast Asian & Western Pacific Region | - | 2 | 1 | 2 | 2 | 5 | 8 | 8 | 8 | 8 | 8 |
| Total | 30 | 40 | 80 | 140 | 130 | 210 | 250 | 180 | 210 | 210 | 220 |

**Table XX: Estimated PDR by UC in millions**

|  | **2030** | **2031** | **2032** | **2033** | **2034** | **2035** | **2036** | **2037** | **2038** | **2039** | **2040** |
| --- | --- | --- | --- | --- | --- | --- | --- | --- | --- | --- | --- |
| UC1 – Delivery in a fixed post by health worker or community health worker | 20 | 30 | 50 | 71 | 74 | 111 | 125 | 100 | 117 | 117 | 117 |
| UC2 - Outreach delivery by health worker | 5 | 7 | 18 | 37 | 36 | 51 | 68 | 46 | 55 | 54 | 58 |
| UC3+UC4 - Outreach or community delivery by community health worker | 7 | 5 | 11 | 28 | 25 | 44 | 52 | 36 | 40 | 43 | 45 |
| Total | 30 | 40 | 80 | 140 | 130 | 210 | 250 | 180 | 210 | 210 | 220 |

**Table XX: Estimated PDR by scenario in millions**

|  | **2030** | **2031** | **2032** | **2033** | **2034** | **2035** | **2036** | **2037** | **2038** | **2039** | **2040** |
| --- | --- | --- | --- | --- | --- | --- | --- | --- | --- | --- | --- |
| Scenario 1 - Base | 30 | 40 | 80 | 140 | 130 | 210 | 250 | 180 | 210 | 210 | 220 |
| Scenario 2 - Regional MR-MAP pilots | 1 | 1 | 30 | 100 | 130 | 150 | 190 | 180 | 210 | 200 | 200 |
| Scenario 3 - Accelerated adoption in countries with greatest need | 120 | 130 | 200 | 210 | 170 | 210 | 230 | 170 | 210 | 200 | 200 |

2.	Gavi The Vaccine Alliance. Measles and measles-rubella vaccine support. Available at: <https://www.gavi.org/types-support/vaccine-support/measles-and-measles-rubella>. Accessed August 13.

8.	United Nations, Department of Economic and Social Affairs, Division P. World Population

Prospects 2019. Geneva, Switzerland, **2019**.

9.	Global Burden of Disease Collaborative Network. Global Burden of Disease study 2019 (GBD 2019) Results. Available at: <http://ghdx.healthdata.org/gbd-results-tool>.

10.	World Health Organization. Immunization Data, vaccine introduction. Available at: <https://immunizationdata.who.int/listing.html?topic=vaccine-intro&location>=. Accessed May.

11.	World Health Organization. Immunization data, provisional measles and rubella data. Available at: <https://immunizationdata.who.int/listing.html?topic=measles-rubella&location>=. Accessed May.

12.	World Health Organization. Immunization expenditure. Available at: <https://immunizationdata.who.int/pages/indicators-by-category/finance.html?ISO_3_CODE=&YEAR>=. Accessed May.

13.	Vynnycky E, Adams EJ, Cutts FT, et al. Using Seroprevalence and Immunisation Coverage Data to Estimate the Global Burden of Congenital Rubella Syndrome, 1996-2010: A Systematic Review. PLoS One **2016**; 11(3): e0149160.

14.	Summary Measles-Rubella Supplementary Immunization Activities, 2000-2020. World Health Organization, **October 2020**.

15.	Rural population. In: Prospects UNPDsWU. New York: World Bank, **2018 Revision**.

16.	Population living in slums (% of urban population). World Bank.

22.	Global Health Workforce Statistics. Geneva: World Health Organization, **2018 update**.
